# Genomics of Acute Myeloid Leukemia at Diagnosis and Remission

**DOI:** 10.64898/2025.12.04.25340141

**Authors:** Kai Yu, Laura W. Dillon, Jesse M. Tettero, Gege Gui, Rasha W. Al-Ali, Michael R. Grunwald, Elizabeth F. Krakow, Elizabeth A. Griffiths, Alexandra Gomez-Arteaga, Rahul S. Vedula, Melhem Solh, Amandeep Salhotra, Nelli Bejanyan, Lori Muffly, Antonio M. Jimenez-Jimenez, Michael W. Drazer, Yi-Bin Chen, Aaron Logan, Reena V. Jayani-Kosarzycki, Sophia R. Balderman, James S. Blachly, Brian C. Shaffer, Lawrence J. Druhan, Cecilia CS. Yeung, Vanessa E. Kennedy, Amir T. Fathi, Hetty E. Carraway, Sandeep Gurbuxani, Melissa Y. Tjota, Farah Sahoo, Max Smith, Dylan Barfield, Jim Guo, James Han, Jason Hu, Heejon Jo, Vidya Kudlingar, Wilfred Li, Yutong Qiu, Pratheesh Sathyan, Sean Truong, Severine Catreux, Sam Ng, Khai Luong, Yunjiao Zhu, Reem Bahr, Jamie Diemer, Christina K. Ferrone, Allison J. Getker, Jackie Bjerke, Stephanie Bo-Subait, Steven M. Devine, Bergetta Dietel, Gabrielle Giammarino, Emily Heying Chihak, Jianqun Kou, Emily Kolb, Danielle O’Donnell Vitali, Stephen R. Spellman, Brenna K. Tesch, Jenny Vogel, Stephanie Waldvogel, Syreeta Weatherspoon, Jeffery J. Auletta, Christopher S. Hourigan

## Abstract

Accurate and comprehensive genetic characterization of acute myeloid leukemia (AML) is essential for diagnosis, prognostication, and treatment selection. We report here, in 255 adults with AML enrolled in a prospective clinical protocol at 18 major cancer centers across the USA, the results of whole genome DNA-sequencing (WGS) at diagnosis and post-treatment remission. WGS effectively recapitulated, and frequently identified genetic alterations missed by, conventional standard of care clinical testing. These new findings included important prognostic and predictive biomarkers, copy number alterations, regulatory element, splicing, and structural variants including partial tandem duplications within KMT2A. All patients had a pathogenic variant detected at diagnosis, and approximately ten percent also had evidence of a potential inherited myeloid malignancy predisposition. This comprehensive atlas of adult AML genomics provides novel insights into disease biology, creates an evidentiary basis to support clinical testing improvements, and is a resource for both diagnostics and drug development. **<This work is embargoed, with agreement of medRxiv, until 7th December 2025>**.

**Statement of Significance:** Acute myeloid leukemia is a diagnostic category encompassing multiple rare hematological malignances. We show, in this nationwide multicenter study, that standardized unbiased whole genome DNA-sequencing and disease-optimized bioinformatics can replicate conventional “standard of care” AML clinical testing results, while also revealing currently underdiagnosed AML disease biology and potential genetic predisposition.

## Main

The importance of genetics in etiology, prognosis, treatment discovery and selection, and monitoring of patients with myeloid leukemias has been increasingly appreciated over the past 50 years. While the first whole genome DNA-sequencing (WGS) of a human cancer 17 years ago was of a patient with acute myeloid leukemia (AML)(1), most of our current evidence regarding adult AML genetics comes from exome(2–4) and targeted panel sequencing approaches(5,6) rather than WGS(2,7). The important associations of AML genomics with prognosis and response to specific treatments are reflected in the recommendations of national and international clinical standard of care guidelines(8,9). In current clinical practice however, there is no harmonized genetic diagnostic testing approach for patients with AML, limiting comparability across datasets and potentially leading to disparities in identifying actionable variants and inconsistent risk assessment based on institutional testing capabilities rather than biological disease characteristics.

AML is a collection of genomically heterogeneous diseases characterized by chromosomal aberrations, gene fusions, and somatic variants(2,3). To address this complexity, the current diagnostic workup has evolved to rely upon a combination of cytogenetic and molecular techniques, including conventional karyotyping, fluorescence in situ hybridization (FISH) and targeted next-generation sequencing (NGS), each designed to detect specific types of alterations. Examples of more comprehensive genomic assessments include optical genome mapping(10), RNA-sequencing(11), and WGS(7) but have not been widely adopted in part due to lack of prospective multicenter comparisons with conventional approaches. Beyond diagnosis and prognostication, increased understanding of AML genomics has also facilitated drug development efforts. Recent regulatory approvals of targeted small molecule inhibitors (Menin(12,13) and mutated *FLT3*(14–16), *IDH1*(17,18), *IDH2*(19)) have ushered in a new era of precision medicine in AML and emphasize the growing need for comprehensive, timely, and high-resolution genomic data in routine practice. WGS offers a compelling alternative to the current inherently fragmented approach where each diagnostic assay captures only a subset of relevant alterations, the specific assay combinations used are inconsistent across centers and even in the same patient over time, and integration of results can be time-consuming, inaccurate, and incomplete.

Here we introduce the AML Temporal Landscape Analysis by Sequencing (“ATLAS”), a nationwide dataset comparing the results of current “standard of care” clinical diagnostic testing for patients with AML evaluated at major academic medical centers in the United States (US) with centralized WGS sequencing performed at diagnosis and remission on a prospective clinical protocol. This resource aims to evaluate the feasibility of WGS as a harmonized and comprehensive replacement for the current multi-platform approach to AML diagnosis, discover new or underappreciated aspects of AML biology with potential prognostic and/or predictive implications, evaluate the utility of additional post-treatment remission testing, and provide a foundational framework supporting future precision medicine initiatives.

## Results

### Cohort overview

Adults with AML in complete remission (CR) were screened at 18 major cancer centers in the US for the prospective multicenter national protocol MEASURE (NCT05224661). Patients were selected for inclusion in ATLAS based on the availability of appropriate stored material from time of AML diagnosis (TOD) and a CR blood sample collected prior to first allogeneic hematopoietic stem cell transplant (alloHCT). Of the 304 patients screened for inclusion, 255 (83.9%) met inclusion criteria for WGS (Supplementary Fig. S1). Patients were enrolled between September 2022 and October 2024. The main reason for exclusion was the lack of appropriate samples.

The median age of patients was 59 years (range 17-79) and 119 (47%) were female (Fig. 1A). Most participants self-identified as White (74%) and of non-Hispanic or Latino descent (87%). Demographic characteristics for patients included were comparable to those patients enrolled in MEASURE but excluded from this ATLAS. TOD samples included bone marrow aspirates in 71% and blood in the remainder. The median time from TOD to CR sample collection was 110 days (range 38-1983). A total of 233 patients (91.0%) were in first complete remission (CR1), while the remaining patients were in second or subsequent CR (CR2+).

**Figure 1:**
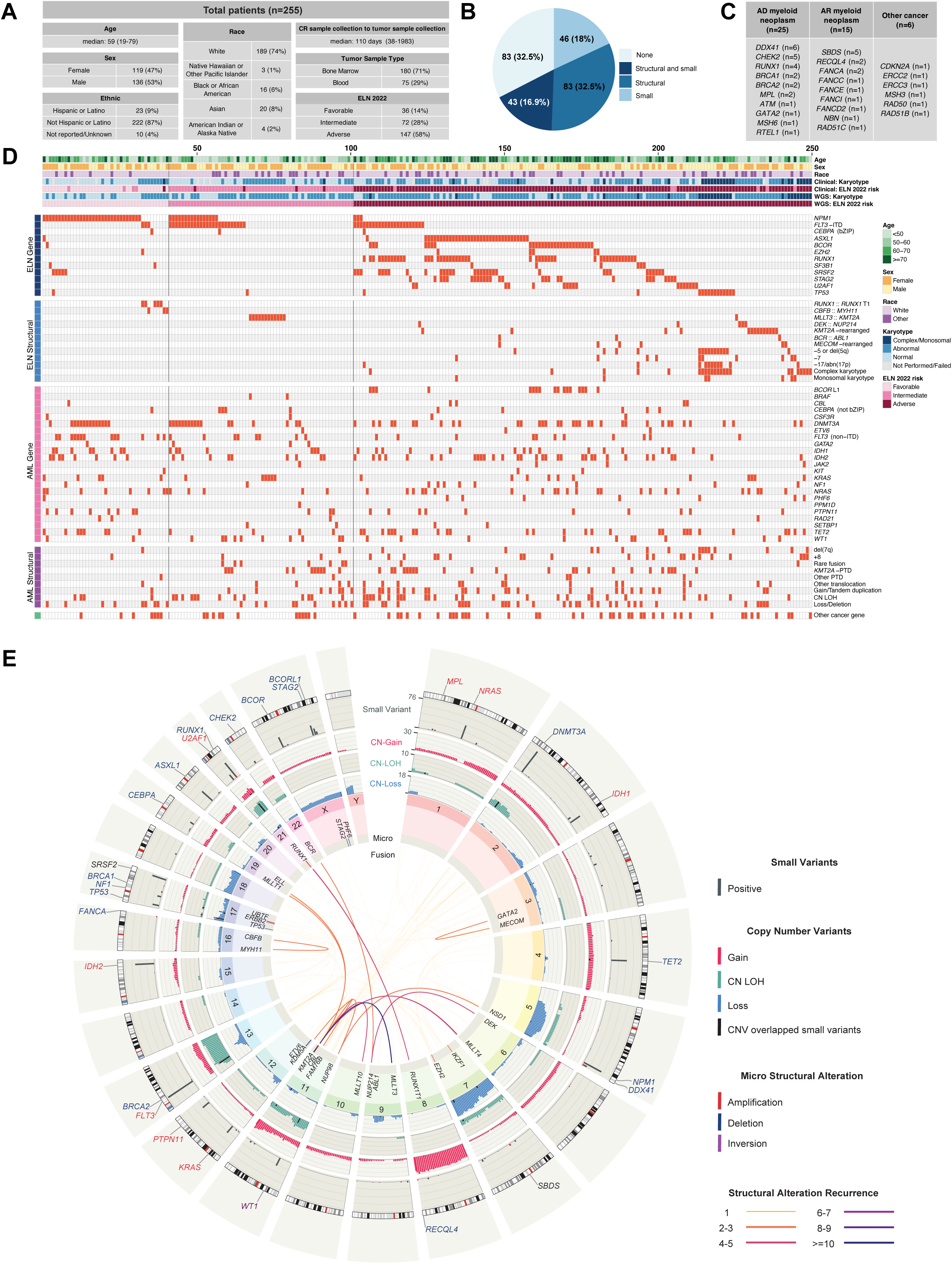
Genomic landscape of ATLAS cohort. **A.** Clinical and demographic characteristics of 255 AML patients undergoing genomic profiling. **B.** The number and percentage of patients without (pale blue, No, n=83) or with novel genomic findings by whole genome sequencing (WGS) not reported by diagnostic clinical testing; including (from light blue to dark blue) small variants (Small, n=46), structural variants (Structural, n=83), or both (Structural and small, n=43). **C.** Potentially germline variants detected by WGS of paired tumor and remission samples. Variants are grouped as autosomal-dominant (AD) and autosomal-recessive (AR) myeloid-predisposition genes as defined by ELN 2022 guidelines and current knowledge for other cancer-susceptibility loci. **D.** Integrated heatmap summarizing baseline clinical characteristics with WGS findings of clinical significance; columns represent individual patients. Baseline clinical characteristics were derived from primary clinical documents and include patient demographics, cytogenetic categories, and derived ELN 2022 risk stratification. WGS results were used to define cytogenetic category, and derived ELN 2022 risk stratification and details are shown as positive (red) for (i) ELN 2022 risk-defining features for AML (ELN Gene, ELN Structural), (ii) extended AML-related features (AML Gene, AML Structural), and (iii) other cancer-related genetic alterations categorized by clinical relevance (Other cancer gene). Partial tandem duplication (PTD), internal tandem duplication (ITD), copy neutral loss of heterozygosity (CN-LOH). Patients with only variants identified in potentially germline genes were removed from this visualization. **E.** Circos plot displays on the cohort level genome-wide distribution of small variants in AML-related genes, copy-number alterations (gain, loss, and CN-LOH), micro-structural variants (<100Kb), and recurrent structural rearrangements. Copy number variants overlapping with a small variant are overlayed in black. Selected cancer-related genes with any potentially germline variant or above 10 somatic variants in this cohort are annotated in the outer ring, while the small variant bars indicating number of patients carried the variant. Text colors indicate: Red: Oncogene; Dark Blue: Tumor suppressor gene; Purple: Oncogene and tumor suppressor gene; Grey: Other.

Documentation from the clinical diagnostic workup performed at the referring centers included complete blood count and leukocyte differential (100%), flow cytometry (97%), cytogenetic (99.2%), and molecular (99.6%) profiling. However, considerable heterogeneity in clinical genetic profiling approaches was observed; patients received a median of 4 (range 1-22) molecular and/or cytogenetic tests (Supplementary Fig. S2A-S2B). The proportion of patients tested using a specific assay type varied widely. Based on centralized review of the clinical testing performed, 36 patients (14.1%) were categorized as favorable risk, 72 (28.2%) as intermediate risk, and 147 (57.7%) as adverse risk according to the 2022 European LeukemiaNet (ELN) classification(8) (Fig. 1A, Supplementary Fig. S2C).

### The genomic landscape of AML revealed by WGS at diagnosis and remission

WGS was performed centrally on TOD samples and blood from CR prior to alloHCT at a median depth of 117X (range 59–300X) and 37X (range 24-89X), respectively (Supplementary Table S1 and S2, Supplementary Fig. S3). In 172 patients (67.5%), WGS identified at least one novel oncogenic or likely oncogenic genomic finding not identified by clinical diagnostic testing, including small variants (46 patients, 18%), structural variants (83 patients, 32.5%), or both (43 patients, 16.9%) (Fig. 1B, Supplementary Table S3 and S4). Additionally, paired TOD and CR sequencing allowed for identification of pathogenic variants of likely germline origin in predisposition genes(20) in 41 patients (16%), including genes associated with autosomal dominant (n=25) and autosomal recessive (n=15) risk of myeloid neoplasms according to ELN 2022 guidelines(8) and the recently described *ATM*(21), as well as other cancer-predisposition genes (n=4 autosomal dominant, n=2 autosomal recessive) (Fig. 1C).

WGS provided information on both small variants and structural variants (copy number gain or loss, gene fusions) necessary for ELN 2022 risk stratification for prognostication or assignment to targeted therapies. WGS also detected alterations missed by standard clinical assessment including small variants in other AML/cancer-related genes and structural variants commonly missed by karyotyping, such as copy-neutral loss of heterozygosity (CN-LOH), micro-structural variants under 5 megabases (Mb), including deletions, and tandem duplications (Fig. 1D-1E).

### WGS can recapitulate standard clinical diagnostic assessment

Following extensive curation and interpretation of WGS variant calls, we identified a median of 4 (range 1-31) genomic events of clinical significance per patient (Fig. 2A). These findings are consistent with prior AML genomics studies (Supplementary Fig. S4).

**Figure 2:**
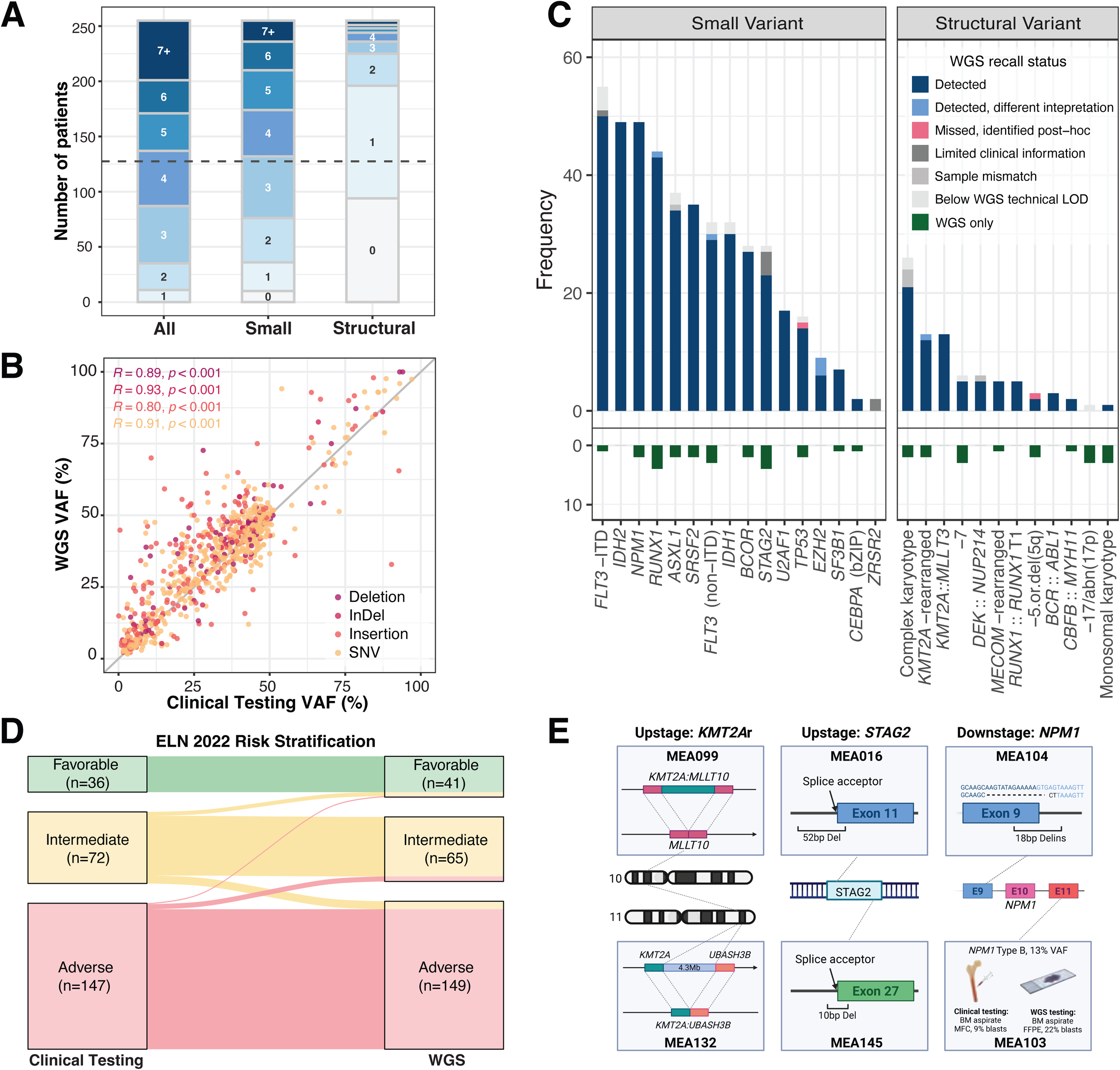
Concordance of tumor-only whole genome sequencing with clinical diagnostic testing and impact on risk stratification. **A.** Frequency distribution of genomic events with clinical significance detected by whole genome sequencing (WGS) per patient. The bars represent the cumulative numbers of patients considering all events (All), molecular events (Small), and structural events (Structural) from left to right, with the dashed line representing the median number of patients. **B.** Correlation of variant allele frequencies (VAFs) between WGS and clinical testing across molecular variant types (*R = 0.80–0.91, p < 0.001*). Each point represents a mutation identified by both methods. **C.** Consistency between WGS and clinical testing for prognostic or predictive genomic features, stratified by molecular or structural abnormality. Bars indicate events detected by WGS (dark blue), detected with alternative interpretation (light blue), missed but identified post-hoc (pink), limited clinical annotation (dark gray), sample mismatch (medium grey), or below the WGS technical limit of detection (light gray). Features only identified by WGS and not by clinical testing are shown in green. **D.** Comparison of ELN 2022 genomic risk stratification based on conventional clinical testing (left) versus WGS (right). Most patients retained the same category (94.5%), whereas 14 (5.5%) were reclassified. Four intermediate risk cases were down-staged to favorable risk, and six were upstaged to adverse risk. Of the adverse risk cases, one was down-staged to favorable and three to intermediate risk. **E.** Representative cases of ELN risk reclassification due to variants called with WGS. Detection of *KMT2A* rearrangements (*KMT2A::MLLT10*, *KMT2A::UBASH3B*) resulted in upstaging to intermediate risk; *STAG2* splice-site deletions led to reclassification to adverse risk; and identification of a novel *NPM1* exon 9/intron 9 Delins and *NPM1* type B insertion undetected by targeted sequencing led to down-staging to favorable risk.

Reporting standards across clinical laboratories are not consistent, making direct comparisons of reported variants with WGS results challenging. However, for well-annotated small variants from the clinical workup, WGS identified the same variants (n=748) with highly correlated variant allele fraction (VAF, r=0.87, p<0.001) (Fig. 2B). For cases where the same sample from clinical testing was used for WGS, 99% of all prognostic and predictive small variants identified clinically were detected by WGS based on the computed WGS limit of detection (LOD) (Supplementary Fig. S5, detailed list in methods). Despite not always having a matched sample or sufficient WGS depth, of the 526 total prognostic and predictive genomic features reported by clinical testing (442 small variants, 84 structural alterations), 489 (93%) were identified independently by WGS, although 6 were interpreted differently by local and central geneticist review (p=0.02, one-sided binomial test for success rate=90%, Fig. 2C). Features reported by clinical testing but not WGS (n=31) were primarily attributable to sample type mismatch (e.g., bone marrow versus blood), incomplete clinical annotation, or variants present below the computed WGS LOD. Conversely, 41 prognostic and predictive genomic features (within 31 patients) were only identified by WGS, 12 of which were not identified in the original clinical evaluation due to a lack of an appropriate test performed and 6 likely due to a sample mismatch.

Application of the ELN 2022 criteria to WGS data yielded identical risk classifications in 237 of 255 patients (93%) (Fig. 2D). Refinement to classification due to additional information from WGS was observed in 11 patients. Eight patients were upstaged to adverse risk, and 3 patients were downstaged to favorable risk. Examples of upstaging and downstaging included identification of cytogenetically cryptic *KMT2A* rearrangements, deletions impacting *STAG2* splice acceptor sites, and identification of *NPM1* variants including a novel insertion deletion variant at the exon 9/intron 9 junction similar to one previously reported to result in NPM1 protein localization in the cytoplasm(22) (Fig. 2E).

### Structural and copy number alterations add to the genomic complexity of AML

WGS enabled the detection of oncogenic variants missed by conventional approaches, including an additional 175 small and 680 structural variant calls compared to clinical testing. New small and structural variants were identified in 49.4% and 41.7% of patients (Fig. 3A, Supplementary Fig. S6). In the 109 patients with cytogenetically normal AML (as defined by a successful normal metaphase karyotype), WGS revealed structural abnormalities in 55 patients (50.5%), including *KMT2A* partial tandem duplication (*KMT2A*-PTD, n=22), CN-LOH (n=20), microdeletions (<5Mb, n=11), cryptic fusions (n=5), copy number gains or losses (n=13), translocations (n=7), and other PTDs (n=2) (Fig. 3B).

**Figure 3:**
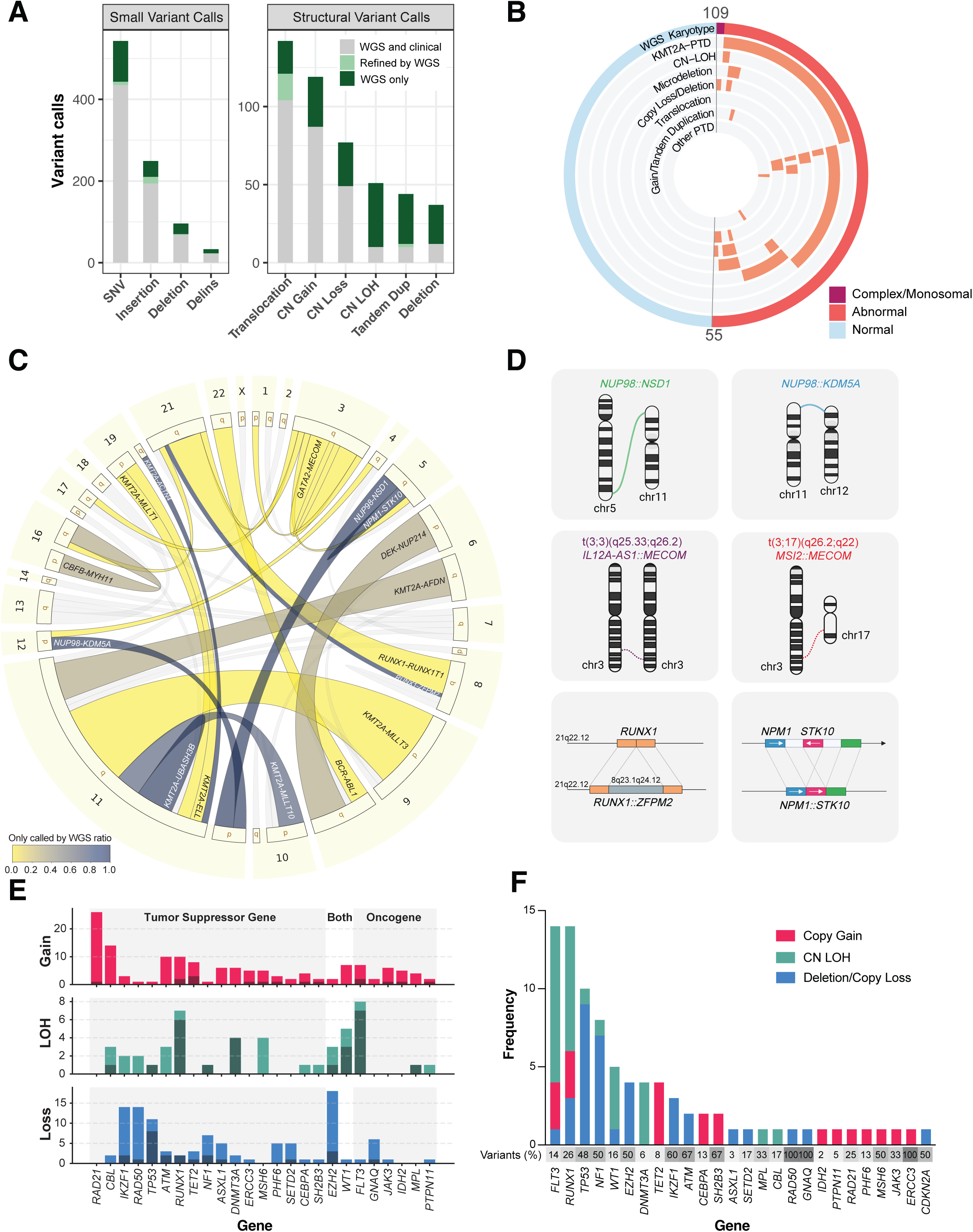
Genome-wide detection of structural and cryptic alterations by whole genome sequencing. **A.** Frequency of small (left) and structural (right) variant calls by whole genomic sequencing (WGS) compared with clinical testing. Bars indicate variants detected by both WGS and clinical testing (grey), refined by WGS (light green), or uniquely identified by WGS (green). **B.** Distribution of WGS-based karyotype (red: complex/monosomal, orange: abnormal, blue: normal) in patients with clinically normal metaphase karyotype (n=109). WGS uncovered additional structural abnormalities (light orange) for 50.5% of the “normal” metaphase karyotype cases (n=55), including *KMT2A* partial tandem duplications (PTD), copy-neutral loss of heterozygosity (CN-LOH), microdeletions (<5Mb), copy gains or loss (>5Mb), translocations, and other PTDs. **C.** Overall recurrence of gene fusion events identified by WGS. The band width represents the number of patients having the fusion event, and the color is scaled by ratio of WGS-identified patients with the fusion event missed by clinical karyotyping and/or FISH assays. **D.** Exemplar cytogenetically cryptic fusion genes identified by WGS. **E.** Distribution of copy number variant (CNV) events (pink: copy gain; green: copy neutral loss of heterozygosity, CN LOH; blue: copy loss) overlapping with AML-related genes and clinically significant small variants within that gene. For each CNV category and gene, the height of each bar represents the number of patients with a CNV event covering the gene; the area marked by a darker shade shows the number of patients who also have a clinically significant small variant in that gene. **F.** Frequency of clinically significant small variants co-occurring with CNV events by gene. The stacked bar plot shows the number of variants co-occurring with CNV events, colored by CNV type. The numbers in the grey boxes represent the percentage of small variants in that gene with a co-occurring CNV event, with darker grey representing higher percentage.

Among all patients, WGS allowed for the identification of driver fusion genes not identified by standard cytogenetics (Fig. 3C), including five *NUP98* rearrangements, which have been associated with sensitivity to *FLT3* inhibitors, venetoclax, and Menin inhibitors(23–25) (Fig. 3D). Atypical *MECOM* rearrangements were also observed, including one t(3;3)(q25.33;q26.2) resulting in *IL12A-AS1::MECOM* and one t(3;17)(q26.2;q22) resulting in *MSI2*::*MECOM*, each having been reported to upregulate *EVI1* expression(26,27). Other cryptic fusions involved *KMT2A*, *RUNX1*, or *NPM1*.

Integration of copy number alteration and small variant data from WGS revealed frequent co-occurrence of pathogenic small variant and copy number alterations in the same gene. For example, CN-LOH involving *FLT3* or *RUNX1* co-occurred with small variants in 7/8 and 6/7 cases, respectively, meanwhile about 50% of *TP53* and *NF1* mutated patients showed co-occurring copy number loss or CN-LOH (Fig. 3E-3F, Supplementary Fig. S7). In all four individuals with potentially germline *RUNX1* variants, somatic CN-LOH events were found that led to the loss of the remaining wild-type allele (Supplementary Fig. S8), suggesting biallelic inactivation during transformation into malignancies.

While the role of extrachromosomal DNA (ecDNA) is increasingly recognized as a mark of genomic plasticity and correlates with oncogene amplification in many cancer types, the occurrence in AML has been reported to be rare(28). In our cohort, ecDNA was detected in 4 cases, involving multiple cancer genes (Supplementary Fig. S9). One patient exhibited *MYC* amplification in a simple circular form, amplified approximately 30 times with *CCDC26*. Three patients had cancer genes (*KMT2A*, *ETS1*, *ERG*, etc.), amplified approximately 5-7 times, all correlated with a complex karyotype.

Complex karyotype is a hallmark adverse genomic feature, but chromothripsis, characterized by focal DNA breakage resulting in copy number alterations and rearrangements, is common in cancer(29) and increasingly appreciated as prognostic in AML(30,31). We observed signatures of chromothripsis in 7 patients, encompassing 30% of those with a complex karyotype (Supplementary Table S5). In all cases, patients had an accompanying oncogenic variant in a DNA damage repair gene known to be associated with chromothripsis, including *TP53* (n=6), and *ATM* (n=1).

These findings highlight the capacity of WGS to uncover cryptic yet clinically actionable structural lesions not detectable by metaphase cytogenetics.

### Enhanced discovery and resolution of clinically relevant genes

In the clinical setting, panel-based molecular tests are commonly used but have a limited capability to detect moderate-sized variants (variants larger than those identifiable by small variant callers but smaller than those identified by metaphase karyotyping). WGS in our study identified diverse subgenic events, including PTDs, internal tandem duplications (ITD), and microdeletions, that were often missed by clinical tests (Fig. 4A). The most frequent was *KMT2A*-PTD, which was detected in 34 cases in 4 forms, most of which (n=24, 71%) were not reported clinically (Fig. 4B, Supplementary Fig. S10A). Interestingly, the location of *KMT2A*-PTD DNA breakpoints was highly enriched within SINE/Alu and LINE elements, suggesting repetitive-sequence mediated recombination (Supplementary Fig. S10B). *RUNX1* microdeletions and PTDs were detected in four patients; both microdeletion and PTD have previously been reported in familial platelet disorder patients as germline alterations(32), but PTD has yet to be reported somatically in myeloid neoplasms (Fig. 4C). Other events found in AML driver genes included microdeletions in *CBL*, *ETV6*, *PHF6*, and *FOXP1,* PTDs in *CCDC26*, *UBTF*, and *EZH2*, and ITDs in *UBTF* and *IKZF1* (Fig. 4A, Supplementary Fig. S10C). Of note, PTDs in lncRNA *CCDC26* were found in 2 patients, which have been suggested to disrupt its normal lncRNA-mediated repression of *KIT*(33).

**Figure 4:**
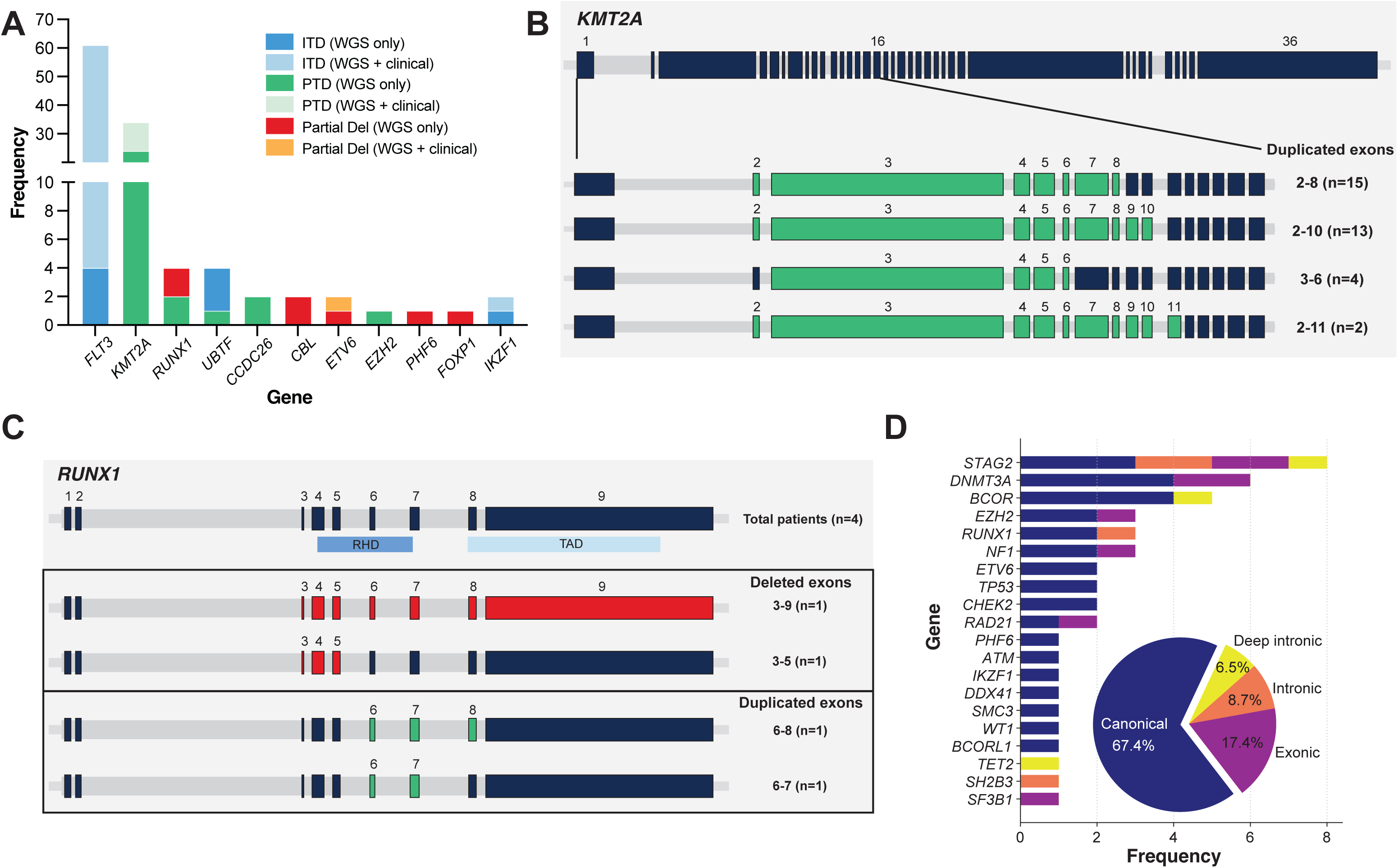
Enhanced gene-level resolution and detection of complex subgenic events by whole genome sequencing. **A.** Frequency of gene-level structural events across the cohort. Stacked bars indicate internal tandem duplications (ITD, blue), partial tandem duplications (PTD, green), and partial deletions (partial del, red). Shading distinguishes events found only by WGS (darker), or both WGS and clinical testing (lighter). The most frequently affected genes were *FLT3*, *KMT2A*, *RUNX1, and UBTF*. **B.** Structural architecture of *KMT2A* partial tandem duplications (PTDs) detected by WGS. Most events involved duplication (green) of exons 2–8 (n = 15) or 2–10 (n = 13); additional configurations (3–6, 2–11) were also observed. **C.** *RUNX1* internal structural events detected by WGS, including deletions (red) spanning exons 3–9 and 3–5 and duplications (green) involving exons 6–8 and 6–7. The diagrams indicate the relative positions of the *RUNX1* runt homology domain (RHD) and transactivation domain (TAD). **D.** Summary of subgenic SpliceAI predicted splicing-altering variants identified by WGS across canonical AML drivers and other important cancer associated genes. The pie chart indicates that 67.4% of detected variants were canonical splice-site variants located in the +1/+2 and −1/−2 bases, whereas 32.6% represented non-canonical or previously unreported configurations, including variants reported as missense or synonymous variants; variants located in intron region that are 2-20 bp away from exon; or deep intronic region (>20bp away from exon) likely to be missed in clinical test panels.

Splice-site altering variants constitute a type of highly clinically relevant but often missed genomic feature. These variants can be far away from canonical splice sites and thus escape detection by targeted or exome sequencing; or exist in non-canonical exon or intron regions and are not easily connected with abnormal splicing(34). We applied SpliceAI prediction(35) on all relevant variants detected in our cohort and identified 46 variants in 20 AML-related genes predicted to be splice-disrupting. More than 30% of the variants were non-canonical (outside the +1/+2 or −1/−2 bases), including 17.4% exonic and 8.7% intronic variants within 20 base pairs (bp) of exon boundaries (Fig. 4D). Importantly, 6.5% were deep intronic variants located >20 bp from exons, which would typically be missed by targeted sequencing. Recurrently affected genes included *STAG2*, *DNMT3A*, *BCOR*, *EZH2*, *RUNX1*, and *NF1*; and deep intronic variants were found in *STAG2*, *BCOR*, and *TET2*.

WGS also allowed for the identification of variants with potential clinical actionability in AML or other diseases that would frequently be missed when performing targeted panel testing specific for myeloid malignancies. ITDs in *UBTF* were observed in 3 cases, which have recently been described as sensitive to Menin inhibitors(36). Loss of function variants in *SETD2* were observed in 5 cases, 4 of which were co-mutated with *KMT2A* (rearranged n=2, PTD n=2), which is associated with cytarabine resistance and could be overcome by combination with S and G2/M checkpoint inhibition(37). Variants in *RIT1* were observed in 5 cases, which have been shown to drive oncogenic transformation in lung tumors and exhibit sensitivity to multiple inhibitors(38). In one case, a truncating variant in *BAP1* was identified, and similar variants have been observed to exhibit sensitivity to PARP inhibition in multiple cancer types(39). Additionally, we observed a patient with a gain of function variant in *SOS1*, an activator of RAS/MAPK pathway, for which multiple inhibitor strategies (including MEK and SOS1) are actively being investigated(40). A total of 3 cases exhibited variants in genes (*POLE*, *LRP1B*, *ARID1A*) with evidence of sensitivity to immune checkpoint inhibitors(41–43).

### Novel genomic features in “dark regions” revealed by WGS

Beyond the common variants directly affecting protein coding, WGS allowed exploratory analysis into the “dark regions” of the human genome to discover potentially relevant genetic features related to AML. Somatic variants in regulatory regions that perturb transcription factor (TF) binding motifs could lead to functional changes related to oncogenesis. In our cohort, mutated motifs with high recurrence included TFs Sp1, PPARa, REST, RUNX3, and EGR2 (Fig. 5A). To screen variants potentially altering gene expression, we applied PromoterAI prediction, combined with PhyloP 30-way conservation score, to highlight that one *TERT* promoter variant could promote its expression level, echoing as a known hotspot with functional consequence in solid tumor(44). Similarly, we discovered five regulatory variants predicted to repress expression in genes *CREBBP*, *DYRK1A*, *ZNF324B*, *FGF11*, and *VAPB* (Fig. 5B). Among these altered genes, *CREBBP* alterations were reported as associated with poor prognosis in *de novo* AML(45); *DYRK1A* expression has been reported as reduced in AML and that overexpression suppresses AML cell proliferation(46); *ZNF324B* has been found to be significantly downregulated in chromothripsis-positive complex karyotype AML patients(31). The motif logo plot indicates that the variants alters the binding sequence of multiple TFs, and CAGE, POLR2A, DNase-seq data support their roles as promoters or enhancers (Fig. 5C, Supplementary Fig. S11A).

**Figure 5:**
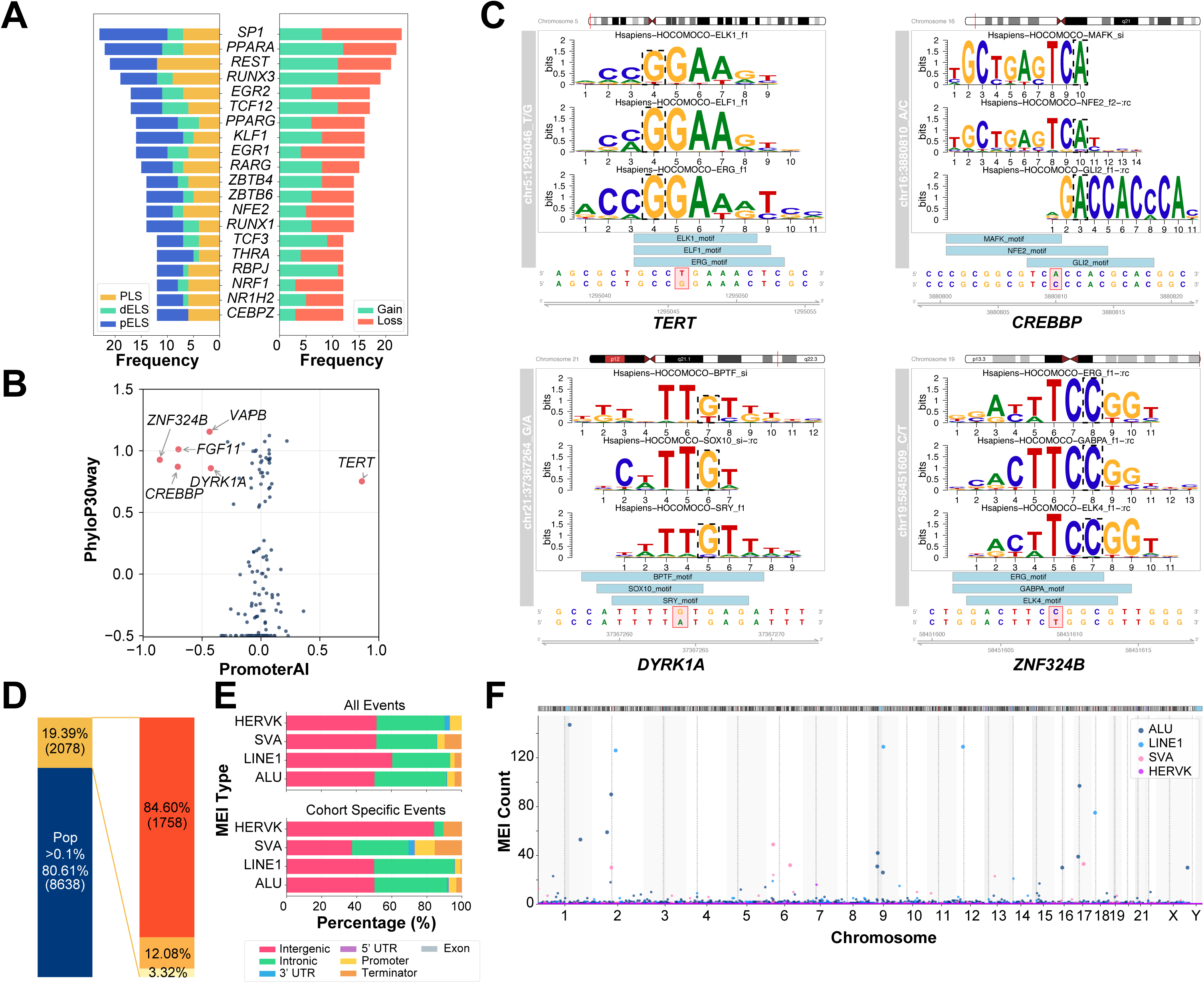
Genome-wide discovery of non-classical genomic alterations by whole-genome sequencing. **A.** Recurrent regulatory-region somatic mutations identified by whole genome sequencing (WGS). Bar colors denote regulatory element type variants (PLS: promoter-like signature, dELS: distal enhancer-like signature, pELS: proximal enhancer-like signature) located (left), and gain or loss of transcription factor binding affinity (right). **B.** Scatterplot of promoter conservation (PhyloP30way) versus predicted impact on gene expression (PromoterAI score), highlighting significantly altered loci. Genes under promotion include *TERT* whereas *ZNF324B*, *CREBBP*, *FGF11*, *VAPB*, and *DYRK1A* show repression. **C.** Representative examples of promoter mutations and their predicted transcription factor–binding changes. Motif disruption or creation is shown for *TERT*, *CREBBP*, *DYRK1A*, and *ZNF324B*, with represented affected motifs. **D.** Composition of Mobile element insertions (MEIs) locus detected in this cohort. The stacked bar plot showed the distribution of detected variants exist in population databases above 0.1% or under 0.1%. The right bar plot breaks down the 2078 MEI locus as “not exist” or “under 0.1%” in the population database. From bottom to top, yellow bar is the number of loci excluded by RepeatMasker simple-repeat and low complex region filter; orange bar is the number of loci excluded by mean allele number and allele number standard deviation filter, left 1758 cohort-specific high-quality loci. **E.** Annotation of MEI integration sites stratified by origin (all events and cohort specific events). Most of the insertions were enriched in intergenic and intronic regions, with a small proportion of events located in promoter and terminator regions. **F.** Chromosomal distribution of cohort specific MEI events. The scattered dots represent MEI counts of each MEI type in 1Mb bins, with different colors representing MEI types.

In our cohort, we identified 10,716 high-quality mobile element insertion (MEI) loci (246,048 events), of which 1,758 (3,476 events) were novel or extremely rare in the general population (population frequency <0.1%, Fig. 5D). With individual level data from 1000 Genomes Project, AML patients exhibited a similar total MEI burden but slightly higher in LINE-1 and SVA types (Supplementary Fig. S11B). For all MEI events identified in this cohort, functional annotation showed that most MEIs were in intergenic or intronic regions, with a small fraction inside promoters or terminators (Fig. 5E). The cohort-specific events had similar distribution except for HERVK, which had a low event number (Supplementary Fig. S11C). When partitioning the genome into 1 Mb windows, the cohort-specific MEIs showed regional enrichment close to the centromere and other loci including 1q31.3, 2p13.2, 2q13, and 6q21 (Fig. 5F, Supplementary Fig. S11D). Compared with CR samples, 84 events were classified as somatic and randomly distributed across the genome (Supplementary Fig. S11E-S11F).

### Integration of diagnostic and remission genomes

WGS profiling of matched CR samples identified both persistent and emergent variants in 114 (44.7%) patients, including both small and structural variants (Fig. 6A, Supplementary Table S6-S7). In total, 107 (42.0%) patients had a persistent somatic variant (VAF ≥2.5% in CR) and 33 (12.9%) patients had an emergent variant (Fig. 6B). Of the 255 patients included in the study, 185 (72.5%) received intensive induction chemotherapy, 66 (25.9%) received less-intensive chemotherapy, and 4 (1.6%) received regimens not classified as either intensive or less-intensive according to the NCCN guidelines(9).

**Figure 6:**
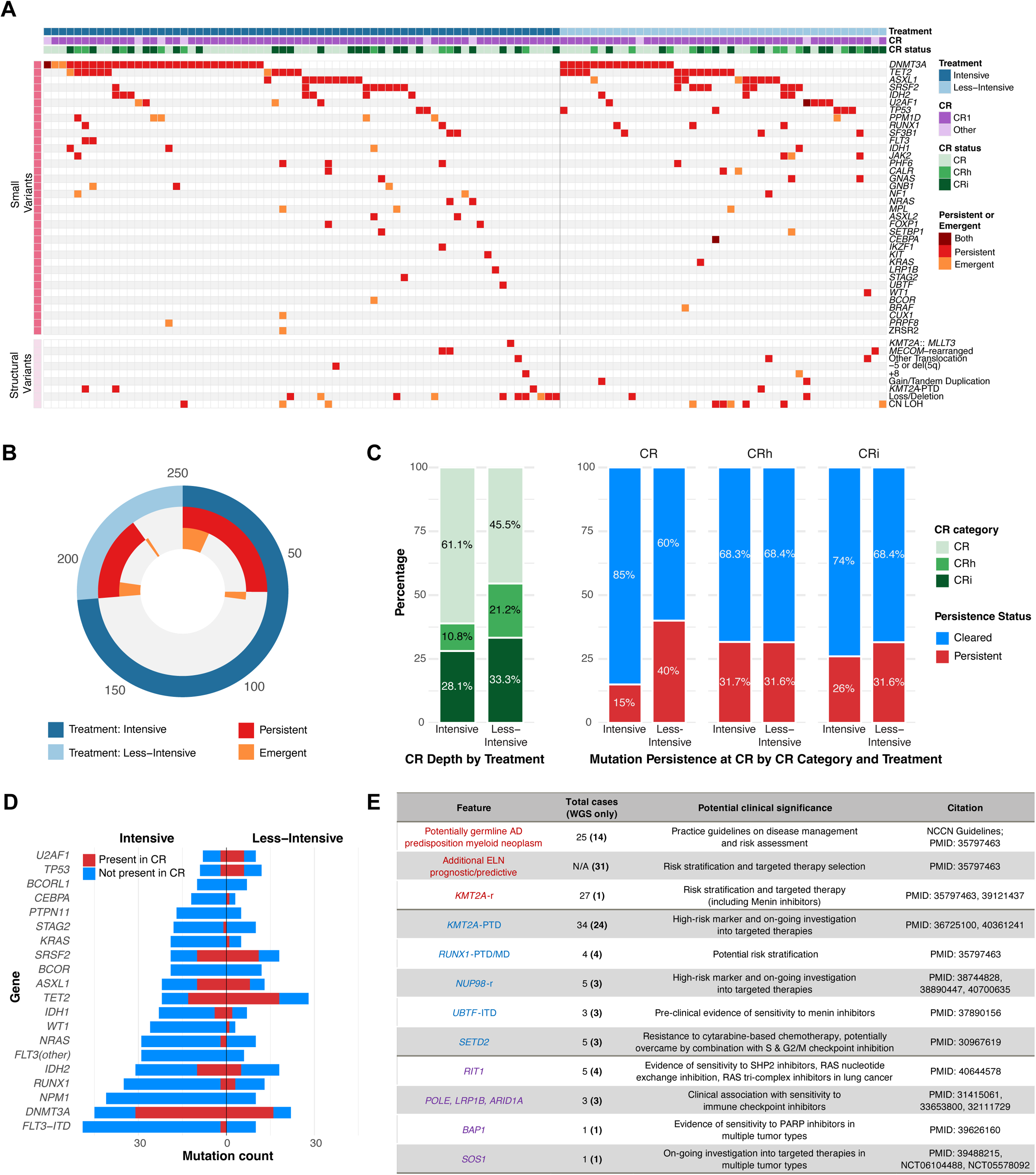
Persistent and emergent variants at complete remission detected by whole genome sequencing. **A.** Heatmap summarizing whole genome sequencing (WGS) profiles of patients with variants of clinical significance in blood collected during complete remission (CR). Genomic features are grouped as small variants or structural variants, ordered by the frequency of total number of patients with events identified in CR. Patients are ordered by treatment groups receiving intensive versus less-intensive treatment. The color indicates features with variants persistent (red), emergent (orange), or both (maroon) at CR. **B.** Distribution of patients with persistent or emergent variants at CR by treatment intensity. **C.** Patient detailed CR status and the association with variant persistence. The left bar plot shows ratios of CR categories, grouped by treatment intensity. The right bar plot shows the variant clearance rate at CR under each CR category, grouped by treatment intensity. **D.** Top 20 recurrently mutated genes at diagnosis and the variant persistence in CR stratified by treatment intensity. Left: variants from patients treated with intensive chemotherapy induction (n=175); right: variants from patients treated with less-intensive regimens (n=64). Persistent variants were more frequent following less-intensive therapy (57.6% vs 33.3%; p<0.001). **E.** Summary of patients with features identified by WGS with potential clinical actionability. The numbers show the total cases (number of patients), with those only identified by WGS in the parentheses. Findings are grouped by evidence level. Red: current guidelines/approved therapies for myeloid malignancies; blue: evidence/on-going trials in myeloid malignancies; purple: evidence/on-going trials in other cancer types. Not applicable (N/A)

The degree of blood count recovery at the CR timepoint differed significantly between intensive (CR 61.1%, CR with incomplete count recovery, CRi, 28.1%, CR with partial hematologic recovery, CRh, 10.8%) and less-intensively treated patients (CR 45.5%, CRi 33.3%, CRh 21.2%), with intensive therapy showing more full CR (p=0.042) (Fig. 6C). For patients in full CR, persistent variants were more often detected in those treated with less-intensive therapy compared to those intensively treated (40% vs 15%, p<0.001), but this difference was not found for patients in CRh or CRi (p=1 and p=0.367, respectively). Variants in *DNMT3A*, *TET2*, *ASXL1*, and *SRSF2* were more commonly seen at initial diagnosis in patients who were subsequently treated with less-intensive induction therapy, and overall, were more likely to persist in remission than being cleared (Fig. 6D). On the patient level, persistent variants in prognostic or predictive genes were more frequent among those treated with less-intensive regimens compared to intensive therapy (57.6% vs 33.3%; p<0.001). Comparison of induction regimens (standard “7+3” [n=93] vs azacitidine-venetoclax [n=43]) revealed distinct post-treatment mutational landscapes (Supplementary Fig. S12). Newly emergent oncogenic variants at CR included *PPM1D* (n=7), *MPL* (n=3), *U2AF1* (n=3), *IDH1* (n=1), and *BRAF* (n=1).

Diagnostic-remission paired WGS enabled the discovery of novel diagnostic features directly correlated with the leukemic portion, including many with prognostic or potentially therapeutic significance (Fig. 6E). Additionally, this analysis shed light on potentially inherited variants and pre-leukemic clonal hematopoiesis; the genome-wide coverage further extended the capability to find events in regulatory or deep intronic regions not captured by standard clinical testing and provides a unique resource for future discovery research.

## Discussion

This ATLAS study has demonstrated that, despite the central importance of genetics in clinical management and drug development for patients with AML, current diagnostic approaches in the US are heterogeneous and can fail to identify important prognostic and potentially actionable features. It has also shown, in the context of a prospective nationwide multicenter clinical protocol, that whole-genome DNA-sequencing is a single, simple, standardized, and rapid test that captures the information currently necessary for clinical practice while also providing important additional insights into underlying disease biology.

In this cohort of 255 adults with AML, relevant prognostic or predictive information was missed in 31 patients (12%) by clinical testing but was detected by WGS. Using WGS, unlike previous reports, no patient with AML was “silent” without an oncogenic variant detected. Additionally, 10% of patients had evidence of a potential autosomal-dominant germline myeloid malignancy predisposition syndrome, a rate analogous to that found in adults with myelodysplastic syndrome undergoing alloHCT(20,47). In our study, over half of those with this potential germline risk would not have been suspected based on the results of the clinical testing performed; suggesting more comprehensive screening may be warranted. Such universal testing would have benefit for patients and their families, and also could add important information during selection of the transplant donor. WGS identified 34 patients (13%) with *KMT2A*-PTD (only 10 of which had been identified clinically). This rate is higher than has been reported using PCR(48,49), targeted NGS panels(50,51), or optical genome mapping(52). While underreporting historically may make prognostic estimates inaccurate, this variant has been associated with high rates of CR attainment, but no clear demonstrated impact on overall survival. Unlike *KMT2A*-rearranged AML (*KMT2A*-r), currently developed Menin inhibition agents alone may not be an optimal therapeutic approach for patients with *KMT2A*-PTD but the high number of observed patients with this genetic feature now motivates both diagnostic test, including measurable residual disease (MRD) testing, and drug development efforts. Identification of *KMT2A*-r, *NUP98*-r, and *UBTF*-ITD, all reported as potentially directly targetable by Menin inhibition, in 9 patients (3.5%) could be more immediately actionable.

There are several limitations to ATLAS. The patients included are all adults with AML that had all achieved CR and were evaluated at a major medical center in the US for consideration of alloHCT. The findings cannot be directly extrapolated to patients with refractory AML, children(53), those considered not suited for alloHCT, or to clinical practice patterns and demographic groups outside the US. Initial AML therapy selection was not randomized, limiting the implications of comparisons between intensive and less-intensive approaches. Patients were enrolled when already in remission, meaning TOD samples were not prospectively collected but instead procured, with patient consent, from previously stored material. This may have created a bias in the patients that enrolled on this study, for example based on referral patterns and site of initial diagnosis, and the nature of the material types available precluded the use of other technology such as RNA-sequencing, single-cell sequencing, or functional studies(3,54,55). Care was taken to select MEASURE clinical sites with geographic diversity across the US. The 69% of non-Hispanic White patients in this initial ATLAS cohort is consistent with the race and ethnicity of recipients of a first alloHCT across the US in the period 2016-2023(56), however the number of patients from other racial and ethnic groups is too small to make meaningful comparisons between groups(57).

Our current prognostic scores and treatment pathways for the care of adults with AML have been based on observed clinical associations with genetic subgroups defined by the technology available historically(8,58). The known wide heterogeneity in clinical outcomes between patients within the same assigned risk group is more understandable in the context of this study, where 50% of normal karyotype patients had structural events. In the full cohort, copy number alterations were common and often associated with oncogenic variants on the other allele in a cancer-associated gene, and both splicing altering and regulatory region variants were undetected using legacy clinical approaches. The patients in this study continue to be followed, and the MEASURE consortium intends to report on clinical outcomes, MRD testing, and relapse biology over the coming years. Collaborations between and with MEASURE consortium investigators are encouraged. A interactive visualization resource for all somatic variants reported in this study will be made available with the peer-reviewed version of this paper. Risk scores are context-specific, and a strength of this study is that it reflects the ongoing change in US clinical practice from intensive cytotoxic chemotherapy to less intensive regimens, in particular hypomethylating agent combination with a Bcl-2 inhibitor(59). The availability of paired WGS from TOD and CR in this resource allows understanding of treatment-specific kinetics of somatic, clonal hematopoietic, and likely germline variants and increased confidence for structural alteration discovery. It also allows additional future exploration of patient specific differences from large reference populations including immunogenetics, mobile element insertion, and other features in the 98% of the human genome not covered by exome sequencing.

Precision medicine for patients with AML will require confident assignment of individuals with similar personal and tumor features as those within the historical evidence. Incomplete genetic profiling may lead to misassignment on a patient level, and ambiguity at a clinical trial or registry level(60). Multiple additional use-cases for WGS beyond AML exist in a clinical diagnostic laboratory; benefits include a technically simple laboratory workflow, the opportunity to standardize across sites and cancer types, speed, scalability, and decreasing costs. While data interpretation was previously a constraint, comprehensive unbiased sequencing with disease-specificity provided by intuitive bioinformatics facilitates rapid clinical reporting. These factors combined now incentivize adoption of WGS as a standard tool in initial AML diagnosis. This nationwide, multicenter study is the most comprehensive genomic analysis of adult AML, captures an associated snapshot of the current clinical diagnostic approach within the US and provides a unique resource for further research and iterative clinical practice improvement.

## Methods

### Ethical approval

The Molecular Evaluation of AML Patients After Stem Cell Transplant to Understand Relapse Events (MEASURE) clinical protocol (ClinicalTrials.gov NCT05224661) was approved by the National Marrow Donor Program (NMDP) Institutional Review Board (IRB). Patients were consented for retrieval and use of banked diagnostic material and for WGS with no results returned directly to patients or their treating physicians. All participants provided written informed consent in accordance with institutional review board-approved protocols and applicable federal and International Council for Harmonization (ICH) Good Clinical Practice (GCP) regulations.

### Patient eligibility

Eligible participants for MEASURE study were adult patients (≥18 years) diagnosed with AML and undergoing alloHCT while in CR. Residual disease detected by MRD testing prior to transplant was not an exclusion criterion. Patients were excluded if they had a diagnosis of acute promyelocytic leukemia or had previously undergone an alloHCT.

The ATLAS cohort included patients enrolled in MEASURE with TOD samples having sufficient DNA for WGS, source documentation of clinical diagnostic workup (including cytogenetic aberrations, variants for ELN disease/risk classification, and additional actionable targets), and evidence that TOD sample had ≥10% tumor burden. Evidence needed to be on matching sample type (bone marrow or blood) within 3 days of TOD sample collection. Acceptable evidence included: flow cytometry; blast percentage by pathology (bone marrow aspirate or complete blood count, as applicable); pathogenic and likely pathogenic variants in myeloid malignancy-associated genes with VAF ≥10%.

### Extraction of clinical testing results

Primary documentation of cytogenetic, molecular, and flow cytometry testing performed during the clinical workup at TOD was uploaded by sites in 21 CFR Part 11 and HIPAA compliant electronic binders for each patient. Documents were reviewed, collated, and ELN 2022 risk stratification assigned by two independent reviewers, then were reviewed in a committee consisting of at least 3 individuals, including a laboratory scientist, medical geneticist, and hematologist/oncologist. Detailed criteria are described in supplementary methods.

### Sample collection

Biological specimens were obtained from consented patients at participating sites, including stored material from TOD and collection of a CR blood sample. In one case, the available TOD sample was from the time of relapse prior to the latest remission rather than initial AML diagnosis. Acceptable sample types included EDTA-anticoagulated whole blood (≥1 mL), viably cryopreserved peripheral blood mononuclear cell aliquots containing ≥1 × 10 cells, bone marrow aspirates (≥1 mL), formalin-fixed paraffin-embedded (FFPE) tissue submitted as blocks or scrolls (≥5 scrolls or a cumulative thickness ≥20 µm), and unstained diagnostic bone marrow smear slides (≥3). All FFPE tissues were confirmed to be non-decalcified prior to processing.

Samples were collected using site-specific standard operating procedures and shipped under temperature-controlled conditions from the sites to the CIBMTR central repository, where they were logged, verified for identity, and stored until requested for analysis. Upon transfer, materials were received and maintained under appropriate storage conditions. Each sample was assigned to a unique laboratory identifier and tracked in the laboratory information management system to ensure complete traceability.

### Sample processing and library preparation

Samples received in primary format were subjected to appropriate DNA extraction workflow in-house, according to the sample type and study timepoint. Genomic DNA, provided by the center or extracted in-house, was first quantified to assign sample appropriate library preparation methodology. Library preparation methodology (including Illumina DNA PCR-free, Kapa HyperPrep PCR-free, NEBNext Ultra II FS DNA PCR-Free and PCR-plus, and NEBNext UltraShear FFPE DNA Library) was selected based on the sample type, DNA quantity, and sequencer availability. Detailed information is described in supplementary methods.

### Genome data analysis

Raw sequencing reads were first processed through an in-house snakemake fastq-to-cram pipeline to evaluate sequencing yield, assess data quality, and rule out any possible sample mix-ups in previous sample collection, processing, and library preparation steps. Pipeline details are described in supplementary methods.

Different from the initial evaluation pipeline, the production workflow for sequencing data processing was performed on TOD and CR samples using the Illumina DRAGEN Heme Whole Genome Sequencing Tumor Only Pipeline (Heme App) (version 4.4.4.62) on the Illumina Connected Analytics (ICA) platform(61). Analysis on ICA was performed on Amazon Web Services (AWS) f2 instances with data also hosted on AWS to maintain compliance with industry-accepted security standards. Paired-end FASTQ files were organized, and the corresponding sample sheets were uploaded to ICA. Samples were processed through the Heme App in batches of 5-10 samples, utilizing default parameters except for a custom bed file as hotspot to enhance the detection of tandem duplications. The hotspot file includes exonic regions of *KMT2A*, *FLT3*, and *UBTF*, and extended by 300 base pairs on both flanking sides, specifically to enhance the detection of ITDs. This pipeline produced BAM files aligned to hg38 reference genome, and small variant, structural variants, and copy number alterations called by DRAGEN (version 4.4.4). All primary variant data were subsequently ingested into Illumina Connected Insights (ICI) (version 5.1.4) for annotation, interpretation, and visualization, resulting in consolidated analytical outputs from the Heme pipeline.

To identify oncogenic variants, variant filters were designed in Illumina Connected Insights to detect structural alterations and small variants in AML-associated genes using a training set of 10 patients. Blinded to clinical test results, a medical geneticist analyzed filtered variants and visual inspection of WGS data in ICA to profile all remaining patients using detailed filter information described in the supplementary methods. After application of filters, variants underwent manual curation to remove any additional artifacts and to extract variants of clinical significance. Variants were assigned oncogenicity following ClinGen/CGC/VICC standards for classification of somatic variants in cancer(62).

Additionally, we applied tumor-CR analysis to call variants in tumor-normal mode, using the Illumina DRAGEN Secondary Analysis Platform (v4.4). Filters were applied to identify additional small structural variants in TOD samples filtered out by the tumor-only pipeline and assigned oncogenticity. Additionally, the output was utilized to identify residual TOD variants in CR below the LOD of tumor-only calling but >2.5% VAF. Additional details provided in the supplementary methods.

Combining the detected variants and their allele-frequency in tumor-only and tumor-CR methods, we identified variants at TOD and CR. Using genes of known association with germline predisposition, variants of potential germline origin were identified and excluded from further analysis of persistence. Finally residual and emerging oncogenic variants at CR were identified. Details on all strategies used for variant identification are described in supplementary methods.

### In-depth genomic feature identification

Additionally, several in-depth analyses were performed to identify additional genomic features, including:

1. Mobile Element Insertion (MEI): Starting from BAM files generated by ICA Heme App, MEIs were identified using MELT (v2.2.2) in “single” mode with the default parameters, using hg38 reference genome and mobile element reference file provided by the software.
2. Splice variant prediction: Starting with all small variants identified by the Heme App, any variant located within 10,000 bp flanking on either side of a transcript were kept for further analysis. The variants were merged into a single variant set with duplicates removed, followed by application of the SpliceAI (v.1.3) prediction algorithm. Parameter (-D 1000) was used for analysis; variants predicted to have any of the four categories (AG, AL, DG, DL) above 0.8 were kept as splicing altering variants.
3. Regulatory element variants: High-confidence somatic variants located in genomic regions annotated by GeneHancer were subject to PromoterAI score calculation. To predict transcription factor binding site (TFBS) disruptions or gains, motifbreakR (v.2.19.5) was applied with the HOCOMOCO motif database. Additional annotation was also extracted including population frequency from gnomAD; evolutionary conservation scores were extracted from UCSC genome browser PhyloP30 database, and regulatory element type from ENCODE cCRE database.
4. Other: AmpliconSuite-pipeline was used to identify ecDNA; ShatterSeek was used to chromothripsis events.

Detailed methods used to generate final results are described in the supplementary methods.

### Statistical analysis

Our hypothesis was that WGS can achieve more than 90% concordance with the clinical workup in identifying abnormalities in the 30 structural and molecular targets listed in “Abnormalities for ELN risk stratification and additional actionable targets”. A one-sided binomial test with 0.05 as the significance level was used. Positive calls of the 30 checkboxes from the clinical workup and the independent WGS workup were extracted for comparison. For patients with a complex karyotype, exact structural alterations were not counted towards the final comparison.

As a secondary analysis, we compared the concordance of two ELN classifications based on clinical and WGS workup respectively. The percentage of concordance in classification were reported for the two methods. For patients with different classifications, identified or unidentified targets using WGS that changed the classification were categorized by assay LOD, new target region, or missing clinical report. For small variants with detailed annotations from clinical workup including VAFs, Pearson correlation coefficient was calculated between the VAFs of variants detected by both methods, with subgroup analyses on detailed variant type.

For the comparison of small variant recall on matched samples, TOD samples used for WGS were required to have the same collection date and sample type (blood or bone marrow) as the clinical reports where we extracted the detailed variant information. The LOD in VAFs given sequencing depth was calculated for each sample based on a linear regression model using experimental data validated down to 5%(63).

## Data availability

An interactive visualization resource for somatic variants and clinical features described here will be published with the peer-reviewed version of the paper. Collaborations with the MEASURE consortium are encouraged and may be proposed to or any MEASURE site principal investigator (https://clinicaltrials.gov/study/NCT05224661).

## Author Contributions

**Conception and design:** C.S. Hourigan

**Development of methodology:** K. Yu, L.W. Dillon, J.M. Tettero, G. Gui, R.W. Al-Ali, C.S. Hourigan

**Acquisition of data (provided animals, acquired and managed patients, provided facilities, etc.):** M.R. Grunwald, E.F. Krakow, E.A. Griffiths, A. Gomez-Arteaga, R.S. Vedula, M. Solh, A. Salhotra, N. Bejanyan, L. Muffly, A.M. Jimenez-Jimenez, M.W. Drazer, Y. Chen, A. Logan, R.V. Jayani-Kosarzycki, S.R. Balderman, J.S. Blachly, B.C. Shaffer, L.J. Druhan, C.C.S. Yeung, P.J. Sung, V.E. Kennedy, A.T. Fathi, H.E. Carraway, S. Gurbuxani, M.Y. Tjota, F. Sahoo, M. Smith

**Analysis and interpretation of data (e.g., statistical analysis, biostatistics, computational analysis):** K. Yu, L.W. Dillon, J.M. Tettero, G. Gui, R.W. Al-Ali, M.R. Grunwald, E.F. Krakow, E.A. Griffiths, A. Gomez-Arteaga, R.S. Vedula, M. Solh, A. Salhotra, N. Bejanyan, L. Muffly, A.M. Jimenez-Jimenez, M.W. Drazer, Y. Chen, A. Logan, R.V. Jayani-Kosarzycki, S.R. Balderman, J.S. Blachly, B.C. Shaffer, L.J. Druhan, C.C.S. Yeung, P.J. Sung, V.E. Kennedy, A.T. Fathi, H.E. Carraway, S. Gurbuxani, M.Y. Tjota, F. Sahoo, M. Smith, D. Barfield, J. Guo, J. Han, J. Hu, H. Jo, V. Kudlingar, W. Li, Y. Qiu, P. Sathyan, S. Truong, S. Catreux, S. Ng, K. Luong, Y. Zhu, R. Bahr, J. Diemer, C.K. Ferrone, A.J. Getker, J. Bjerke, S. Bo-Subait, S.M. Devine, B. Dietel, G. Giammarino, E. Heying Chihak, J. Kou, E. Kolb, D. O’Donnell Vitali, S.R. Spellman, B.K. Tesch, J. Vogel, S. Waldvogel, S. Weatherspoon, J.J. Auletta, C.S. Hourigan

**Writing, review, and/or revision of the manuscript:** K. Yu, L.W. Dillon, J.M. Tettero, G. Gui, R.W. Al-Ali, M.R. Grunwald, E.F. Krakow, E.A. Griffiths, A. Gomez-Arteaga, R.S. Vedula, M. Solh, A. Salhotra, N. Bejanyan, L. Muffly, A.M. Jimenez-Jimenez, M.W. Drazer, Y. Chen, A. Logan, R.V. Jayani-Kosarzycki, S.R. Balderman, J.S. Blachly, B.C. Shaffer, L.J. Druhan, C.C.S. Yeung, P.J. Sung, V.E. Kennedy, A.T. Fathi, H.E. Carraway, S. Gurbuxani, M.Y. Tjota, F. Sahoo, M. Smith, D. Barfield, J. Guo, J. Han, J. Hu, H. Jo, V. Kudlingar, W. Li, Y. Qiu, P. Sathyan, S. Truong, S. Catreux, S. Ng, K. Luong, Y. Zhu, R. Bahr, J. Diemer, C.K. Ferrone, A.J. Getker, J. Bjerke, S. Bo-Subait, S.M. Devine, B. Dietel, G. Giammarino, E. Heying Chihak, J. Kou, E. Kolb, D. O’Donnell Vitali, S.R. Spellman, B.K. Tesch, J. Vogel, S. Waldvogel, S. Weatherspoon, J.J. Auletta, C.S. Hourigan

**Administrative, technical, or material support (i.e., reporting or organizing data, constructing databases):** K. Yu, L.W. Dillon, J.M. Tettero, G. Gui, R.W. Al-Ali, J. Bjerke, S. Bo-Subait, S.M. Devine, B. Dietel, G. Giammarino, E. Heying Chihak, J. Kou, E. Kolb, D. O’Donnell Vitali, S.R. Spellman, B.K. Tesch, J. Vogel, S. Waldvogel, S. Weatherspoon, J.J. Auletta, C.S. Hourigan

**Study supervision:** C.S. Hourigan

## Acknowledgements

This work was supported by the Office of Naval Research, the Intramural Research Program of the National Heart, Lung, and Blood Institute, and the Red Gates Foundation and was a research collaboration with Illumina Inc.

Sequencing was performed, in part, at the NHLBI Intramural DNA Sequencing and Genomics Core. This work utilized the computational resources of the NIH HPC Biowulf cluster (https://hpc.nih.gov/) and the Advanced Research Computing at Virginia Tech (https://arc.vt.edu/).

The CIBMTR is a research collaboration between the Medical College of Wisconsin and the NMDP and is supported primarily by Public Health Service U24CA076518 from the National Cancer Institute (NCI), the National Heart, Lung and Blood Institute (NHLBI) and the National Institute of Allergy and Infectious Diseases (NIAID); 75R60222C00011 from the Health Resources and Services Administration (HRSA); N00014-24-1-2057 and N00014-25-2146 from the Office of Naval Research.

The views expressed in this article do not reflect the official policy or position of the National Institute of Health, the Department of the Navy, the Department of Defense, Health Resources and Services Administration (HRSA) or any other agency of the U.S. Government.

Thanks to our colleagues Dr. Marc Schwartz, Dr. Pamela Sung, and Dr. Wael Saber for their involvement in the clinical protocol and Dr. Gloria Garcia and Emma Frasez Sørensen for helpful feedback on the preparation of this manuscript.

