## Supplementary Methods for "Genomics of Acute Myeloid Leukemia at Diagnosis and Remission"

### 1. Sample processing

DNA extraction was performed according to the sample type and study timepoint, as described below.

#### 1.1 DNA Extraction from time of diagnosis peripheral blood or bone marrow

At diagnosis (baseline AML), genomic DNA was extracted from various sources including: formalin-fixed paraffin-embedded (FFPE) tissue sections, peripheral blood or bone marrow smear slides, and cryopreserved peripheral blood or bone marrow mononuclear cells. FFPE-derived DNA was isolated using the QIAamp DNA FFPE Advanced UNG Kit (Qiagen, cat#56604), per the manufacturer's recommendations, incorporating deparaffinization, proteinase digestion, and DNA purification. DNA from smear slides and cryopreserved cells was extracted using the Quick-DNA™ Miniprep Plus Kit (Zymo Research, cat# D4069). All extractions were performed under standardized conditions to ensure DNA purity and integrity while minimizing cross-contamination.

Eluted DNA was quantified using the Qubit™ 4 Fluorometer with the Qubit™ 1X dsDNA High Sensitivity Assay Kit (Thermo Fisher, cat#Q33230). Concentration and yield data were recorded in the laboratory information management system, and each sample was assigned a unique internal ID for traceability.

#### 1.2 DNA extraction from remission peripheral blood

Genomic DNA was isolated from 1 mL of peripheral blood collected in EDTA tubes using the chemagic *360* instrument (Revvity) with the chemagic DNA Blood H24 Kit (cat. CMG-1097). Up to 24 samples were processed per batch, with manifests generated for traceability. DNA was eluted in 300 µL of IDTE buffer (10 mM Tris, 0.1 mM EDTA, pH 8.0, Integrated DNA Technologies, cat#11-05-01-13) and quantified using the Quant-iT™ PicoGreen™ dsDNA Assay Kit (Invitrogen, P7589) on the BioTek Cytation 7 (Agilent). DNA concentration and yield data were recorded in the laboratory information management system, and each sample was assigned a unique internal ID for traceability.

### 2. WGS library preparation and sequencing

Genomic DNA, provided by the center or extracted in-house, was first quantified to assign sample appropriate library preparation methodology. Library preparation methodology was selected based on the sample type, DNA quantity, and sequencer availability.

In general, for high quality gDNA extracted from fresh or flash frozen bone marrow and blood samples, PCR-free methods were prioritized to acquire a more uniform representation of the genome. In the early phase, 275-350 ng high quality genomic DNA samples were submitted to Broad Clinical Laboratories for PCR-free WGS service, using Kapa Biosciences HyperPrep library construction kit (KK8515). When genomic DNA yield was lower than 275ng but greater than 100ng, the samples were assigned to an in-house PCR-free workflow, using NEBNext Ultra II FS DNA PCR-Free Library Prep Kit for Illumina (E7430L) to prepare libraries with 100–150 ng input gDNA. In these cases, we followed the default library preparation protocol and used an 8-minute enzymatic fragmentation time to generate libraries with a targeted average insert size of approximately 450 bp. For the majority of our cohort, libraries were generated using the Illumina DNA PCR-Free Prep, Tagmentation (20041795) kit, with 300 ng of input DNA. For samples with limited material, the workflow was successfully adapted to use as little as 100 ng input without compromising sequencing success.

Due to different sample availability, for samples with less than 100 ng high quality genomic DNA, PCR amplification was required to produce enough library for sequencing. We employed the NEBNext Ultra™ II FS DNA Library Prep Kit (E7805L) for in-house PCR library preparation. In this workflow, we also used an 8-minute fragmentation time to achieve a 450 bp target size. PCR cycles were adjusted according to manufacturer recommendations based on DNA input mass, balancing the need for sufficient library with minimizing amplification-induced artifacts.

For the challenging FFPE-derived genomic DNA samples, which often have lower extraction yield as well as fragmented and chemically modified DNA, we applied in-house FFPE WGS workflow using NEBNext UltraShear FFPE DNA Library Prep Kit (E6655S). Based on the sample availability, 10-250ng FFPE gDNA were repaired using the NEBNext FFPE Repair Mix v2 following default protocol. A 15 min enzymatic fragmentation and limited PCR amplification were done following manufacturer's recommendation to maximize library preparation efficiency.

For all library types, dual index adapters were used to avoid index hopping during sequencing. For PCR-free, PCR amplified normal and FFPE libraries, NEBNext Multiplex Oligos for Illumina (Unique Dual Index UMI Adaptors DNA Set 1, E7395S) was used for library preparation. For PCR-free libraries made with Illumina DNA PCR-Free Prep kit, IDT for Illumina - DNA/RNA UD Indexes Set A were used. For PCR-free libraries made by Broad Clinical Laboratory, custom Broad indices were used.

For all in-house WGS libraries, their concentration was measured using either Qubit™ ssDNA Assay Kit (Illumina DNA PCR-free Prep) or QuantStudio 5 qPCR machine with KAPA Library Quantification Kits (KK4824). PCR-free library size was estimated based on fragmentation conditions used in the experiment. To get the fragment size of PCR amplified libraries, we performed a quality control check on Agilent 4200 TapeStation System with High Sensitivity D1000 ScreenTape.

The majority of libraries were prepared and sequenced in house, with Illumina NovaSeq 6000 and NovaSeq X systems, targeting 120X depth for TOD samples and 30X depth for CR samples. A subset of sample libraries were sequenced at Broad Clinical Labs with NovaSeq X plus system, targeting 60X depth for TOD samples and 30X depth for CR samples.

### 3. Genome data analysis

#### 3.1 Data quality control

The raw sequencing data were first processed through an in-house snakemake(1) fastq-to-cram pipeline on local high-performance computational cluster to evaluate sequencing yield, assess data quality, and rule out any possible sample mix-ups in previous sample collection, processing, and library preparation steps.

In brief, low-quality bases were removed using fastp(2) (v.0.24.0, default parameters, with optional use of -t 1 -T 1 to trim the terminal base of all reads), also data quality metrics were collected for quality control. The cleaned reads were aligned to hg38 reference genome using bwa with optional parameter (-K 10000000), then processed with GATK(3) (v.4.5.0.0) in multiple steps: 1. SortSam (by coordinate); 2. MarkDuplicates (--OPTICAL_DUPLICATE_PIXEL_DISTANCE 2500); 3. GATK Base Quality Score Recalibration (BQSR) workflow, which corrects systematic biases in base quality scores by modeling error rates relative to known variant sites. The post-BQSR BAM files were merged to sample level and converted to CRAM format by samtools(4) (v.1.19.2), to reduce storage footprint. To collect sequencing depth and coverage statistics information, we used GATK CollectWgsMetrics to estimate the genome coverage, informing possible top-up sequencing needed for any samples not reaching the aimed depth.

To rule out potential sample swaps, we employed the fingerprinting strategy by extract fingerprint genotype information using GATK ExtractFingerprint against a recommended panel of common SNPs from the GATK resource bundle (Homo_sapiens_assembly38.haplotype_database.txt). the sample level fingerprint VCFs were then cross-compared across the cohort, using GATK CrosscheckFingerprints (--CROSSCHECK_BY READGROUP --NUM_THREADS 1 --CALCULATE_TUMOR_AWARE_RESULTS true). In this setting, we evaluated the cross-contamination at both sample and read group level. All comparison results indicated that no unexpected matches were detected in our dataset.

#### 3.2 Variant filters applied on Illumina DRAGEN Heme WGS Tumor Only (HemeApp) result

1. Variant category: small variant
    a. Genes: *ABL1, ANKRD26, ASXL1, ASXL2, ATM, ATRX, BCOR, BCORL1, BRAF, CALR, CBL, CBLB, CCDC26, CCND1, CCND2, CCND3, CDC25C, CDKN2A, CEBPA, CHEK2, CSF3R, CUX1, CXCR4, DDX41, DNMT3A, ETNK1, ETV6, EZH2, FANCL, FBXW7, FLT3, FOXP1, GATA1, GATA2, GNAS, GNB1, HRAS, IDH1, IDH2, IKZF1, JAK2, JAK3, KDM6A, KIT, KMT2A, KRAS, LUC7L2, MAP2K1, MPL, MYC, MYD88, NF1, NOTCH1, NPM1, NRAS, PDGFRA, PDGFRB, PHF6, PIGA, PPM1D, PRPF8, PTEN, PTPN11, RAD21, RB1, RIT1, RUNX1, SAMD9, SAMD9L, SETBP1, SETD2, SF3B1, SH2B3, SMC1A, SMC3, SRSF2, STAG2, STAT3, STAT5B, SUZ12, TET2, TP53, U2AF1, U2AF2, UBTF, WT1, ZRSR2*
        i. Frequency in gnomAD <=0.05
        ii. Deleterious consequence (start loss, stop gained, stop loss, incomplete terminal codon, feature elongation, feature truncation, splice donor variant, splice acceptor variant, splice region variant, frameshift variant, inframe deletion, inframe insertion, missense variant, protein altering variant, coding sequence variant)
        iii. VCF filter: PASS
    b. Gene: *NPM1*
        i. Consequence: frameshift variant
        ii. Genomic region (hg38): chr5:171410527-171410565
        iii. Allele depth >=2

  c. Gene: *FLT3*
    i. Consequence: inframe insertion
      1. Genomic region (hg38): chr13:28033878-28034298
      2. Allele depth >=2
    ii. Variant type: Insertion, Delins
      1. Genomic region (hg38): chr13:28033878-28034298
      2. Allele depth >=2
  d. Gene: *UBTF*
    i. Consequence: inframe deletion, inframe insertion
      1. VCF filter: PASS
  e. Gene: any
    i. VCF filter: pass
    ii. Frequency in gnomAD <=0.05
    iii. Actionability: OncoKB(5): Level 1, Level 2, Level 3A, Level 3B, Level R1, Level Dx1, Level Dx2, Level Dx3, Level Px1, Level Px2, Level Px3; CKB(6): Tier 1A, Tier 1B, Tier 2C
2. Variant category: structural variant
  a. Variant type: RNA fusion variant
    i. Split read count >=2, paired end read count >=2
    ii. Gene pairs:
      1. *RARA* with *STAT5B*, *NPM1*, *FIP1L1*, *TBL1XR1*, *ZBTB16*, *STAT3*, *IRF2BP2*, *BCOR*, or *PML*
      2. *RUNX1* with *RUNX1T1*
      3. *CBFB* with *MYH11*
      4. *DEK* with *NUP214*
      5. *PRDM16* with *RPN1*
      6. *RBM15* with *MRTFA*
      7. *NPM1* with *MLF1*
      8. *NUP98* with *NSD1* or *KDM5A*
      9. *ETV6* with *MNX1*
      10. *KAT6A* with *CREBBP*
      11. *PICALM* with *MLLT10*
      12. *FUS* with *ERG*
      13. *RUNX1* with *CBFA2T3*
      14. *CBFA2T3* with *GLIS2*
      15. *BCR* with *ABL1*
      16. *KMT2A*, *MECOM*, *NUP98*, or *ETV6* with any gene partner and VCF filter: PASS
  b. Variant type: structural variant, copy number variant
    i. VCF filter: pass
    ii. Variant length >=5Mbp
  c. Variant type: structural variant, RNA fusion variant
    i. VCF filter: pass
    ii. *MECOM*: genomic region (hg38): chr3:167895854-169895865
    iii. *GATA2*/*RPN1*: genomic region (hg38): chr3:128264999-128845375
    iv. *RARA*: genomic region (hg38): chr17:40331179-40348453

d. Variant type: insertion, tandem duplication
    i. Consequence: feature elongation
    ii. Supporting reads >=2
    iii. *FLT3*: genomic region (hg38): chr13:28033878-28034298
e. Variant type: tandem duplication
    i. VCF filter: PASS
    ii. Gene: *CCDC26*, *UBTF*
f. Variant type: copy number variant
    i. VCF filter: PASS
    ii. Gene: *MYC*, *ERBB2*
    iii. CNV fold change >=2

#### 3.3 Illumina DRAGEN Tumor-CR analysis

Somatic variant calling was performed using the Illumina DRAGEN Secondary Analysis Platform (v4.4.4) in tumor-normal mode. Input FASTQ files from tumor and matched CR samples were processed through the DRAGEN germline small variant caller pipeline to generate BAM and germline small variant VCF files, followed by somatic variant detection through the DRAGEN small variants, structural variants, and CNV pipelines. Small variants (SNVs and indels) were identified using DRAGEN's haplotype-based somatic caller, enabling sensitive detection of low-frequency variants. The somatic caller employs sample-specific noise models and a joint Bayesian analysis of tumor and normal samples to robustly distinguish true somatic mutations from germline variants and sequencing artifacts(7). To enhance the filtering of possible germline variant leakage, variants identified as population germline are annotated in the INFO field with 'GermlineStatus', leveraging data from population databases such as the 1000 Genomes Project and gnomAD(8). Structural variants were detected using DRAGEN SV caller, a next-generation algorithm that builds on and significantly advances the underlying Manta framework by leveraging breakpoint-graph assembly, multi-genome mapping, and machine-learning-based scoring for markedly improved sensitivity and breakpoint resolution(9). Copy number variations were called using DRAGEN's CNV module, which applies a shifting-levels model and incorporates read-depth and discordant read-pair signals to resolve allele-specific copy number changes and loss-of-heterozygosity events(9). To further enhance the accuracy of CNV detection, both the germline-aware and VAF-aware modes were enabled. The germline-aware mode (https://help.dragen.illumina.com/product-guide/dragen-v4.4/dragen-dna-pipeline/cnv-calling/available-pipelines/somatic-cnv-calling-wgs#germline-aware-mode) uses germline copy number information to adjust the expected read depth contribution from normal cell contamination in the tumor sample. This process enables more accurate assignment of somatic copy numbers in regions affected by germline CNVs. The VAF-aware mode (https://help.dragen.illumina.com/product-guide/dragen-v4.4/dragen-dna-pipeline/cnv-calling/available-pipelines/somatic-cnv-calling-wgs#vaf-aware-mode) evaluates the proportion of reads supporting each allele, which helps resolve complex copy number changes and increases confidence in identifying allele-specific events, such as loss of heterozygosity or subclonal amplifications.

All analyses were restricted to a high-confidence genomic BED file derived from the Genome in a Bottle (GIAB) genome stratification resource(10), specifically the "notinalldifficultregions" category from the GRCh38 v3.1 stratifications (https://github.com/genome-in-a-bottle/genome-stratifications/blob/master/GRCh38/v3.1-GRCh38-all-stratifications.tsv).

All somatic variant calls were generated in standard VCF format, with each variant further annotated using Illumina's Nirvana engine(11) through the Illumina Connected Analytics (ICA) platform. DRAGEN's somatic SNV and indel caller has demonstrated high sensitivity and precision relative to other methods, while delivering results in significantly less time. This speed advantage is enabled by DRAGEN's unified, hardware-accelerated pipeline, which provides a scalable and reproducible workflow for high-throughput somatic variant analysis(7).

Step-by-step example commands used for the Tumor-CR analysis are provided below:

Step1: Mapping and germline small variant calling

```
dragen -f \
--validate-pangenome-reference false \
 --ref-dir <path-to-hg38-alt_masked.graph.cnv.hla.rna_v3> \
--output-directory <output-path-directory> \
--output-file-prefix <sample-name> \
--fastq-list <path-to-sample-fastqlist> \
--enable-map-align true \
--enable-map-align-output true \
--enable-duplicate-marking true \
--enable-variant-caller true
```

Step2: Tumor-CR somatic small variant and structural variant calling

```
dragen -f \
--enable-map-align false \
--enable-map-align-output false \
--ref-dir <path-to-hg38-alt_masked.graph.cnv.hla.rna_v3> \
--output-directory <output-path-directory> \
--output-file-prefix <sample-name>  \
--bam-input <CR_sample_BAM_path> \
--tumor-bam-input <TOD_sample_BAM_path> \
--enable-variant-caller true \
--enable-variant-annotation true \
--variant-annotation-data </path/to/your/NirvanaData> \
--vc-enable-germline-tagging true \
--enable-sv true
```

Step3: Germline-aware and VAF-aware CNV calling

3a: Germline CNV calling from CR sample

```
dragen -f \
--ref-dir <path-to-hg38-alt_masked.graph.cnv.hla.rna_v3> \
--output-directory <output-path-directory> \
--output-file-prefix <CR-sample-name>  \
--bam-input <CR_sample_BAM_path> \
```

```
--enable-metrics-json true \
--enable-map-align false \
--enable-cnv true \
--cnv-enable-self-normalization true
```

3b: Somatic CNV calling from TOD sample

```
dragen -f \
--ref-dir <path-to-hg38-alt_masked.graph.cnv.hla.rna_v3> \
--output-directory <output-path-directory> \
--output-file-prefix <TOD-sample-name>  \
--tumor-bam-input <TOD_sample_BAM_path>  \
--enable-map-align false \
--enable-cnv true \
--cnv-enable-ref-calls true \
--cnv-enable-self-normalization true \
--cnv-normal-b-allele-vcf <CR_germline_snv_vcf> \
--cnv-somatic-vc-vcf <TOD/CR_somatic_snv_vcf> \
--cnv-normal-cnv-vcf <CR_germline_cnv_vcf>
```

#### 3.4 Germline predisposition variant identification

To identify variants of potential germline origin associated with a predisposition to developing myeloid malignancies or other cancer types, we utilized the results from paired TOD and CR WGS, with the theory that variants of germline origin would be present at high VAF (>30%) in both the TOD and CR samples. The output from the Illumina DRAGEN Tumor Only Heme App from TOD and CR was filtered in ICI as follows:

1. Variant type: Small variant
    a. Gene: any
    b. VCF filter: PASS
    c. Frequency in gnomAD <=0.05
    d. Deleterious consequence (start loss, stop gained, stop loss, incomplete terminal codon, feature elongation, feature truncation, splice donor variant, splice acceptor variant, splice region variant, frameshift variant, inframe deletion, inframe insertion, missense variant, protein altering variant, coding sequence variant, gain of function variant, loss of function variant, switch of function variant)
    e. VAF >= 0.30 (at TOD and CR)

Remaining variants interpreted as oncogenic/likely oncogenic were investigated for potential germline predisposition consequence as follows:

1. Hematologic malignancies (based on ELN 2022 guidelines and known literature)
   Gene: *ATM, BRCA1, BRCA2, MLH1, MSH2, MSH6, PALB2, PMS2, TP53, CEBPA, DDX41, RUNX1, ANKRD26, ETV6, GATA2, ELANE, G6PC3GFI1, HAX1, JAGN, TCRG1, VPS45A, SBDS, DNAJC21, EFL1, SRP54, FANCA, FANCB, FANCC, FANCD2, FANCE, FANCF, FANCG, FANCI, FANCJ, BRIP1, FANCL, FANCM, RAD51C, SLX4, ERCC4, RAD51, UBE2T,*

*XRCC2, REV7, RFWD3, FAP100, ACD, CTC1, WRAP53, RPA1, DCLRE1B, DKC1, MDM4, RTEL1, TERC, TERT, TINF2, NHP2, NOP10, NPM1, PARN, CBL, PTPN11, NRAS, KRAS, NF1, SAMD9, SAMD9L, BLM, CHEK2, MPL, RECQL4, NBN, WAS*

2. Other cancer predisposition (based on ACMG secondary findings and known literature)
   Gene: *APC, BMPR1A, MAX, MEN1, MUTYH, NF2, PTEN, RB1, RET, SDHAF2, SDHB, SDHC, SDHD, SMAD4, STK11, TMEM127, TSC1, TSC2, VHL, WT1, CDKN2A, ERCC2, ERCC3, MSH3, RAD50, RAD51B*

#### 3.5 Somatic variant identification

Somatic variants were identified by integrating information collected from tumor-only analysis of the TOD and CR timepoints, as well as the paired Tumor-CR analysis. Multiple strategies were employed.

1. Somatic variants identified at TOD and CR using tumor-only analysis.
   a. Filters as described in the "Variant filters applied on Illumina DRAGEN Heme WGS Tumor Only (Heme App) result" section applied to the TOD and CR outputs separately.
   b. Variants identified and interpreted as oncogenic/likely oncogenic were curated independently.
   c. Variants of potential germline origin described above were removed.
   d. A variant was determined to be a residual somatic variant in CR if it was identified in both the TOD and CR samples.
      i. For copy number variants, passing CR variants were manually inspected for overlap with tumor calls.
   e. A variant was determined to be an emergent somatic variant in CR if it was not identified in the TOD sample.

Several limitations in comparing tumor and CR samples arise from using Heme App single sample analysis, and therefore integration of results from the paired tumor-CR analysis were employed.

To balance specificity, sensitivity, and analysis speed, the tumor-only Heme App analysis was fine-tuned to only reports structural variants > 50Kb (if located outside of the defined hotspot regions), which could lead to failure to detect some intra-chromosomal events of oncogenic significance. To capture mid-sized structural variants that are larger than those called by the small variant filter but smaller than those called by the tumor-only structural variant caller, *de novo* discovery from tumor-CR was performed.

2. Somatic mid-sized SVs identified at TOD using paired TOD-CR analysis:
   a. For SVs identified in paired tumor-CR analysis, we retained only deletions, insertions, and inversions, and excluded any events larger than 5 Mb.
   b. SVs were required to meet the following criteria:
      i. FILTER = PASS or MinSomaticScore present, to account for variants potentially not cleared at CR
      ii. Overlapping genomic regions of AML-related genes
      iii. TOD VAF > 5%
      iv. Passing SVs were subjected to manual inspection to remove low-quality or artifactual calls and compared with tumor-only SV calls to avoid duplicate events reported.

v. The remaining variants were interpreted for oncogenicity.

Additionally, since somatic variants could be retained in the CR sample at a level below the LOD of 30X WGS tumor-only analysis, we incorporated results from Tumor-CR analysis to improve detection of low frequency residual variants

3. Somatic residual variants in CR using Tumor-CR analysis:
    a. Variant identified as oncogenic/likely oncogenic by tumor-only analysis of TOD sample.
    b. The variant is detected in CR with a VAF >= 2.5% (equivalent to heterozygous variants using 5% blast criteria for a clinical CR).
    c. The variant is detected in CR in at least 2 alternate reads (to reduce the chance of a false positive call due to PCR or sequencing error).

**3.6 In-house analysis modules**

*3.6.1 Mobile Element Insertion (MEI) Analysis*

Starting from BAM files generated by Heme App, MEIs were identified using MELT(12) (v2.2.2) in Single mode using the default parameters, using hg38 reference genome and mobile element reference file provided by the software. The output VCF files were annotated with AnnotSV(13) using the parameters (-annotationMode full -genomeBuild GRCh38 -includeCI 1 -overwrite 1 -SVinputInfo 1 -SVminSize 50 -overlap 70 -snvIndelFiles -snvIndelPASS 1).

High quality MEI calls were filtered according to strategies previously described(14). Events without low-complexity tag, passed VCF filters, supported by more than 2 split reads, and ASSESS value ≥3 were retained.

To evaluate population frequencies, MEI events were first compared with gnomAD structural variation database(15) (v4.1). All MEI events with ALT field annotated as any type of "<INS:ME>" were included. Matches to gnomAD were categorized into four types: 1. Exact: identical genomic location and MEI type; 2. Overlap: above 80% of the event length overlapped with a gnomAD MEI item as the same MEI type; 3. exact_generic: identical genomic location, however the MEI type was not annotated by gnomAD; 4. overlap_generic: MEI event matches 80% of the event length, however the MEI type was not annotated by gnomAD. For events matching any of these four categories, gnomAD match type, location, MEI type, population maximum allele frequency (popmax) information was extracted. In addition to gnomAD, population frequency information was also extracted from the SV database of high-coverage 1000 Genome Project(16), and the subset of Human Mobile Element Insertion Database(14) that is called from 2677 1KGP samples.

Because genome coverage variability could lead to underreporting MEIs in population database in some genome regions, gnomAD all sites allele numbers were extracted at each MEI locus, including 25 bp flanking regions on both sides. The mean allele number in population and the standard deviation of the allele number within each region were calculated. MEI loci were filtered out if their allele number standard deviation > 10,000 or mean allele number < 50,000. The MEI loci that are annotated by RepeatMasker as simple repeat or low complexity were also removed.

Somatic MEI events were identified by comparing the post-population-frequency-filters events with their paired CR sample. For each high-quality MEI in the tumor sample, a matching CR event was defined if an identical MEI type was reported at the same location, or within the distance of 1/10 of the MEI length (up to a maximum of 50 bp). If no matching MEI was found in the CR sample, we try to extract soft-clip events from BAM file. If at least 3 reads soft-clipped at aligned location within a 50 bp window, this soft-clip event was considered as a matched CR event. VAF and depth was calculated based on the total and soft-clipped read count. Tumor-CR matched events were excluded if any of the following applied: tumor depth < 30; CR depth < 10; tumor VAF < 0.1; tumor VAF was not above 5 times higher than CR VAF. The retained MEIs were classified as somatic events.

#### *3.6.2 SpliceAI score calculation*

With Heme App identified small variants in each patient, all variants located in a region of any transcript with 10,000 bp flanking on both sides were kept for SpliceAI(17) analysis. The variants were merged into a single variant set and removed duplicates, then split into multiple trunks to apply SpliceAI (v.1.3) analysis in parallel between multiple GPU nodes. Parameter (-D 1000) were used for analysis, variants predicted to have any of the four categories (AG, AL, DG, DL) above 0.8 were kept as splicing altering variants.

#### *3.6.3 PromoterAI score calculation*

Variant calls from Illumina Tumor-CR analysis pipeline were first filtered to include high-confidence somatic variants. Variants were retained if: (1)Tumor AF > 10%; (2)Tumor depth > 30; (3) Normal depth > 10; (4)Normal AF < 10%; (5)Tumor VAF / Normal VAF > 5. Filtered VCF files from all patients were subsequently merged using bcftools(4) (v.1.9) to generate a cohort-level variant dataset. Focusing on regulatory regions, variants were then restricted to genomic regions annotated by GeneHancer(18) (v.5.24).

For each retained variant, their variant consequence, population frequency was annotated with Nirvana. ENCODE cCRE database(19) (v.4) was used to annotate each variant to different categories, including promoter-like signatures (PLS), proximal enhancer-like sequences (pELS), and distal enhancer-like sequences (dELS). Their associated target genes were extracted from GeneHancer database(18). Additionally, evolutionary conservation scores were extracted from UCSC genome browser PhyloP30 database(20). To predict transcription factor binding site (TFBS) disruptions or gains, motifbreakR(21) (v.2.19.5) were applied with the HOCOMOCO motif database(22) (v.11).

To evaluate the potential functional impact on regulatory activity, PromoterAI scores were used in this analysis. First of all, precomputed scores were extracted for variants that were covered. For unannotated sites, local prediction scores were generated with PromoterAI(23) (v.1.0rc6) on a GPU cluster.

With all annotated fields described, variants and their associated motif changes were further refined through a multi-step filtering process. For all variants motifbreakR annotated as strong effects, the variant will be excluded if it absent from ENCODE cCRE database; or lack of bone marrow hematopoietic tissue annotation from GeneHancer; or GeneHancer connected_score < 300; or gnomAD popmax > 0.1%; or variant annotated as coding variant; or the transcription factor gene expression level (nTPM) < 1 (The Human Protein Atlas(24) version 19.3 and Ensembl version 92.38).

*3.6.4 ecDNA identification*

BAM files produced by the Heme App were used to identify focal amplification and extrachromosomal DNA (ecDNA). In brief, AmpliconSuite-pipeline(25,26) (v.1.3.9) were used in this study. The pipeline was executed with GRCh38 reference and paired tumor and CR sample BAM files from each patient. The analysis activated both AmpliconClassifier and AmpliconArchitect module with default parameters. The visualization of detected ecDNA were done using CycleViz (v.0.2.2)

*3.6.5 Chromothripsis events detection*

To detect chromothripsis events, we used ShatterSeek(27) to analyze each patient's CNV and SV data. CNV segments and SV events were extracted from Illumina Heme App-produced CNV and SV VCF files with PASS VCF filters and converted to the required format. Chromothripsis calls were identified using suggested filter considerations and manually inspected for confirmation.

### 4. Clinical data analysis

#### 4.1 ELN 2022 classification

Risk stratification of each patient was performed based on the European LeukemiaNet (ELN) 2022 guidelines on risk stratification of AML in adults(28). A risk classification was assigned based on (1) documents provided from testing performed by the clinical site and (2) variants identified by WGS. The assignment of risk stratification was reviewed by a committee consisting of at least 3 individuals, including a laboratory scientist, medical geneticist, and hematologist/oncologist. As treatment intensity information was not available at the time of classification, all patients were provisionally categorized under the intensive treatment protocol.

Abnormalities for ELN risk stratification and additional actionable targets include 30 in total.

1. ELN risk stratification
   a. Structural abnormalities:
      - t(9;11)(p21.3;q23.3)/*KMT2A::MLLT3*
      - t(v;11q23.3)/*KMT2A*-rearranged
      - t(9;22)(q34.1;q11.2)/*BCR*::*ABL1*
      - t(8;21)(q22;q22.1)/*RUNX1*::*RUNX1T1*
      - inv(16)(p13.1q22) or t(16;16)(p13.1;q22)/*CBFB*::*MYH11*
      - t(6;9)(p23.3;q34.1)/*DEK*::*NUP214*
      - t(8;16)(p11.2;p13.3)/*KAT6A*::*CREBBP*
      - inv(3)(q21.3q26.2) or t(3;3)(q21.3;q26.2)/*GATA2*, *MECOM*(*EVI1*) or t(3q26.2;v)/*MECOM*(*EVI1*)-rearranged
      - t(15;17)(q24;q21)/*PML*::*RARA*
      - −5 or del(5q)
      - −7
      - −17/abn(17p)

- Complex karyotype
- Monosomal karyotype

b. ELN molecular targets

- *NPM1*
- *FLT3*-ITD
- bZIP in-frame mutated *CEBPA*
- *ASXL1*
- *BCOR*
- *EZH2*
- *RUNX1*
- *SF3B1*
- *SRSF2*
- *STAG2*
- *U2AF1*
- *ZRSR2*
- *TP53*

2. Other actionable targets

- *IDH1*
- *IDH2*
- *FLT3*-TKD

#### 4.2 Treatment Classification

Patient treatment was categorized as intensive or less-intensive according to the *NCCN Clinical Practice Guidelines in Oncology: Acute Myeloid Leukemia*(29). Intensive regimens included CLAG-M, CLIA, CPX-351, FLAG-IDA, HiDAC, and variants of the standard *"7+3"* induction using either daunorubicin or idarubicin. Less-intensive regimens comprised of azacitidine monotherapy, azacitidine + ivosidenib, azacitidine + venetoclax, decitabine + venetoclax, cladribine + low-dose cytarabine (LDAC), cladribine + LDAC + venetoclax, and LDAC monotherapy.

### 5. Other data analysis framework and data Illustration

R version 4.4.0, including several R packages, was used for statistical tests and visualization(30-32). Pandas(33) have been used as data structural during analysis. Tools including matplotlib(34) and pycirclize(35) were used for data illustration. Part of the model plots were created with BioRender.com and ProteinPaint(36).
