## Supplementary Figures for "Genomics of Acute Myeloid Leukemia at Diagnosis and Remission"

A

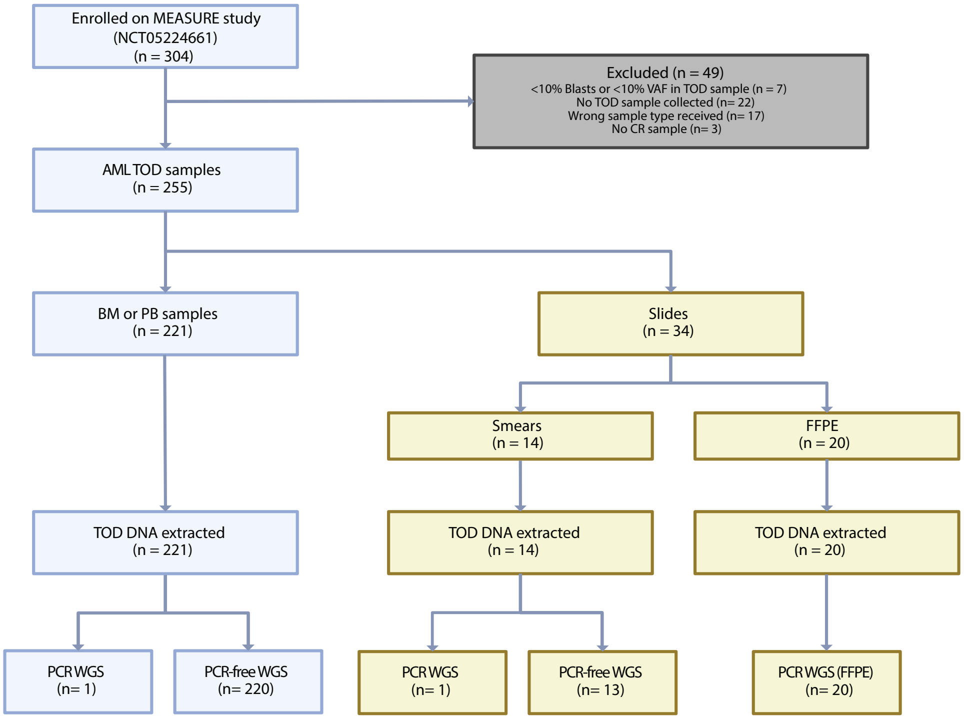


B

| | **ATLAS** | **Excluded** | **Total** |
|---|---|---|---|
| **Age**, median (range) | 59 (19-79) | 58 (27-77) | 59 (19-79) |
| **Sex** | | | |
| Female | 119 (47%) | 25 (51%) | 144 (47%) |
| Male | 136 (53%) | 24 (49%) | 160 (53%) |
| **Ethnic** | | | |
| Hispanic or Latino | 23 (9%) | 4 (8%) | 27 (9%) |
| Not Hispanic or Latino | 222 (87%) | 44 (90%) | 266 (87%) |
| Not reported/Unknown | 10 (4%) | 1 (2%) | 11 (4%) |
| **Race** | | | |
| White | 189 (74%) | 41 (84%) | 230 (76%) |
| Native Hawaiian or Other Pacific Islander | 3 (1%) | 0 (0%) | 3 (1%) |
| Black or African American | 16 (6%) | 3 (6%) | 19 (6%) |
| Asian | 20 (8%) | 2 (4%) | 22 (7%) |
| American Indian or Alaska Native | 4 (2%) | 1 (2%) | 5 (2%) |

**Supplementary Fig. S1. Consort diagram (A) and MEASURE patient demographics (B).**

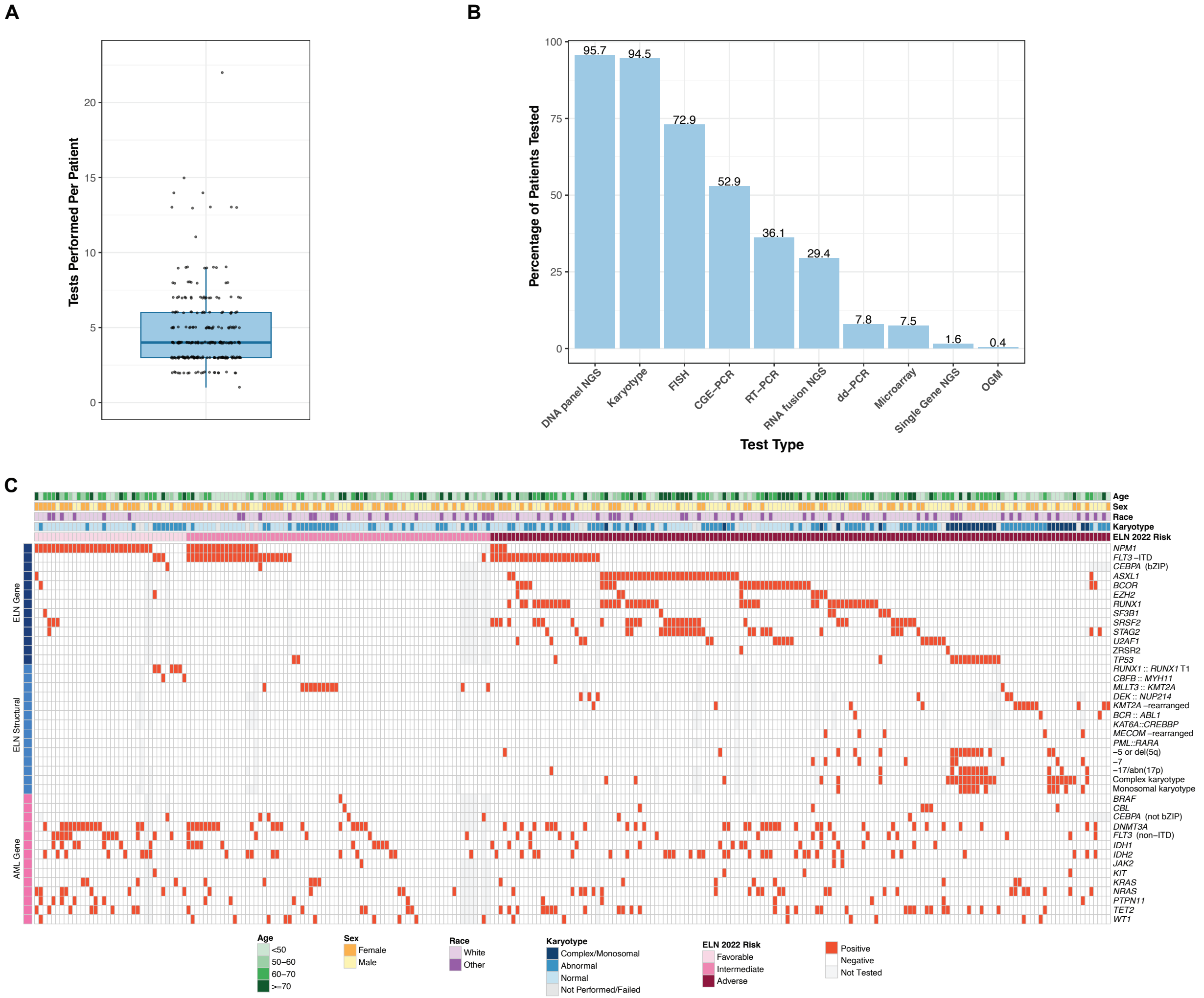


**Supplementary Fig. S2. Summary of clinical diagnostic genetic testing. (A)** Patients received a median of 4 (range 1-22) molecular and/or cytogenetic tests. **(B)** The proportion of patients tested using a specific assay type varied widely. Data were compiled from primary documentation of cytogenetic and molecular testing at each site. See *Methods*, "Extraction of clinical testing results," for details. **(C)** Heatmap summarizes baseline clinical characteristics and genomic features of clinical significance reported by clinical genetic testing. Baseline clinical characteristics include patient demographics (age, sex, race), cytogenetic category, and derived ELN 2022 risk stratification. Details on clinical testing results are shown as positive (red) for: small variants in prognostic/predictive genes (ELN Gene), risk defining structural alterations (ELN Structural), and extended AML-related features (AML Gene).

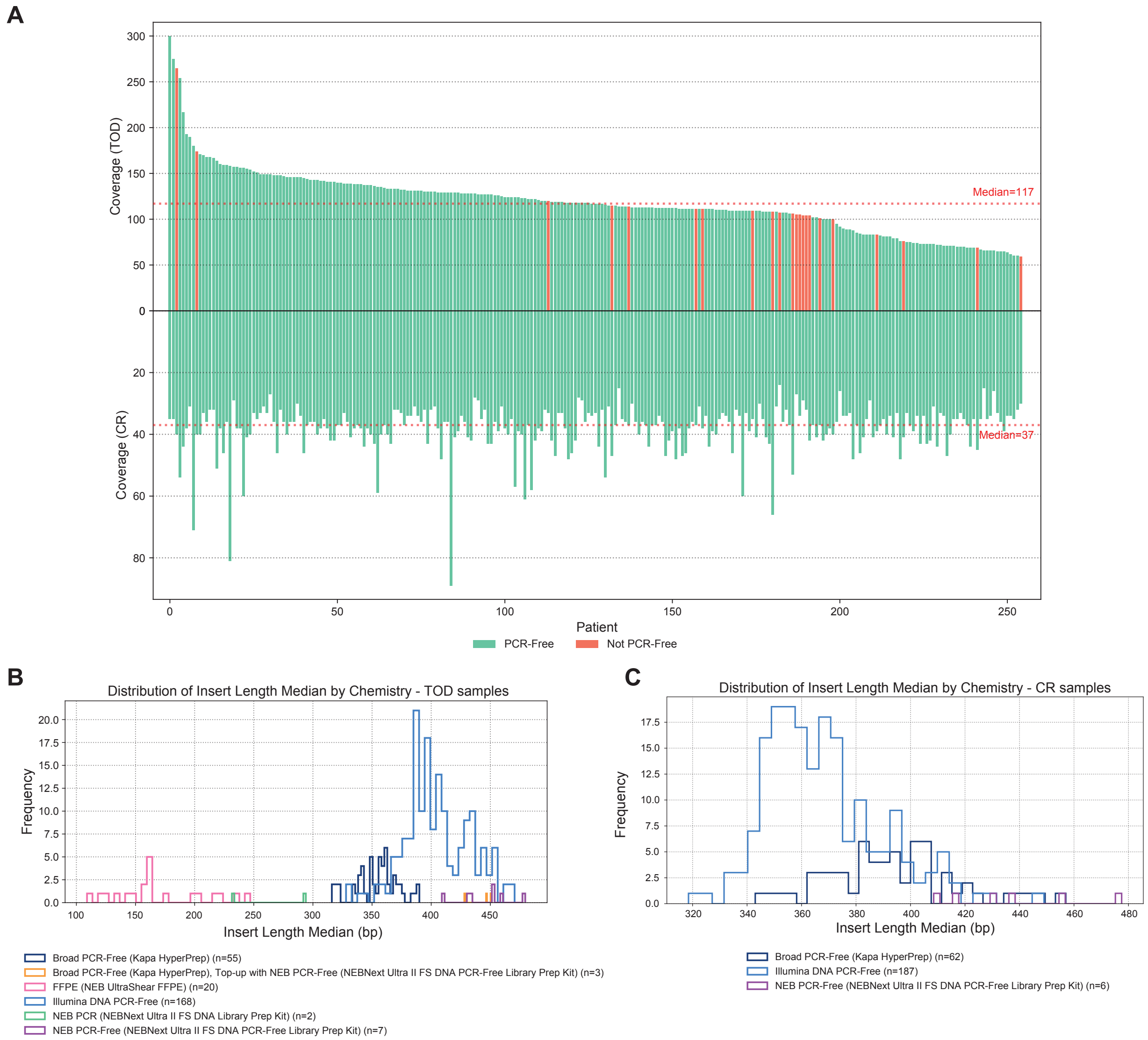


**Supplementary Fig. S3. Summary of WGS run metrics across ATLAS cohort. (A)** Barplot demonstrating average autosomal coverage of Time of Diagnosis (TOD, top panel) and Complete Remission (CR, bottom panel) samples for each patient. Patients are ordered by TOD coverage and bars are colored by PCR-Free status of the libraries. **(B)** Distribution of library insert size across TOD samples, grouped by library preparation chemistry. **(C)** Distribution of library insert size across CR samples, grouped by library preparation chemistry.

**A**

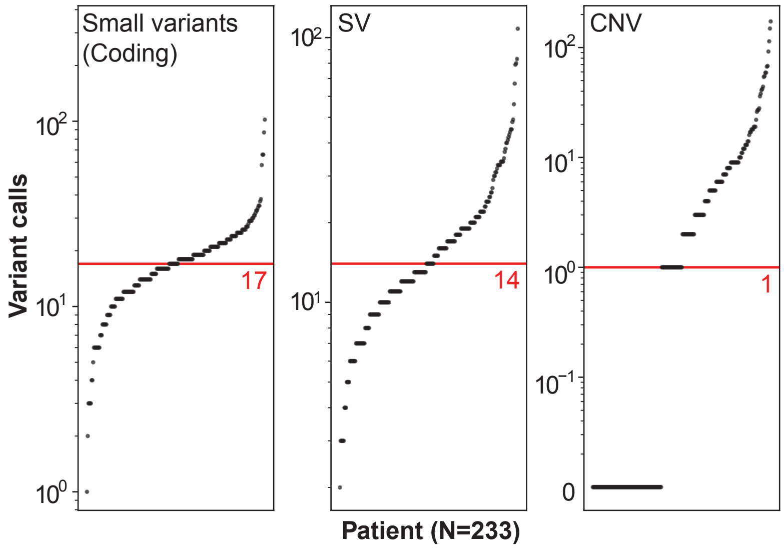


**B**

| Study | Cohort size | Analytical platform | Somatic small variants | Genomic events of clinical significance |
|---|---|---|---|---|
| The Cancer Genome Atlas PMID: 23634996 | 200 adult AML | WGS or WES | Average 13 (range 0-51) variants in coding genomic regions/patient | N/A |
| BeatAML PMID: 30333627 | 531 AML | WES | Median 13 (range 1-80) filtered variants/patient | N/A |
| Papaemmanuil *et al* PMID: 27276561 | 1540 adult AML | 111 gene panel + karyotype | N/A | 86% of patients with ≥2 mutations, 5234 driver mutations in 1540 patients = ~3.4/patient |
| Duncavage *et al* PMID: 33704937 | 263 patients, 175 AML | WGS | N/A | Mean 5.1 (range 0-20) mutations/patient |
| Current study | 255 adult AML | WGS | Median 17 (range 1-102) variants in coding genomic regions/patient | Median 4 (range 1-31) mutations/patient |

**Supplementary Fig. S4. Distribution of variants identified by whole genome sequencing (WGS) at time of diagnosis and overview of variant metrics from previous AML genomic studies. (A)** For patients with PCR-free WGS performed (n=233), a median of 17 (range 1-102) small variants in coding genomic regions, 14 (range 2-108) structural variants (SV), and 1 (range 0-173) copy number variants (CNV) were observed. **(B)** Comparison of high-level variant metrics between the current study and other AML genomic studies.

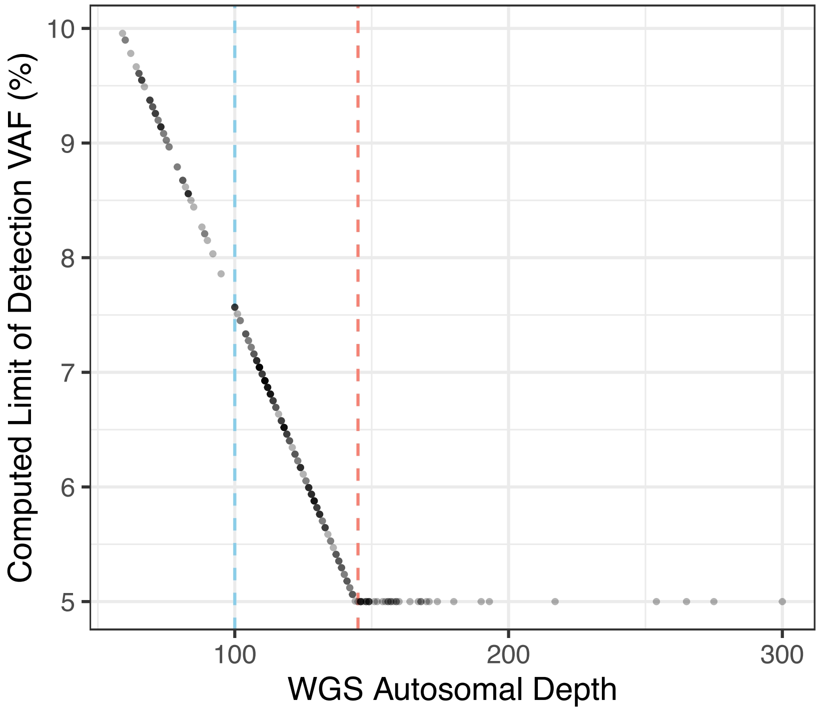


**Supplementary Fig. S5. Computed whole genome sequencing (WGS) limit of detection (LOD) per case based on autosomal sequencing depth.** The LOD of variant allele frequency (VAF) was based on a linear regression model from Illumina's experimental data, which were validated down to 5%. The red dashed line denotes the depth required for the detection of variants with a VAF ≥5%, and the blue dashed line denotes the median WGS autosomal sequencing depth for the cohort at time of diagnosis.

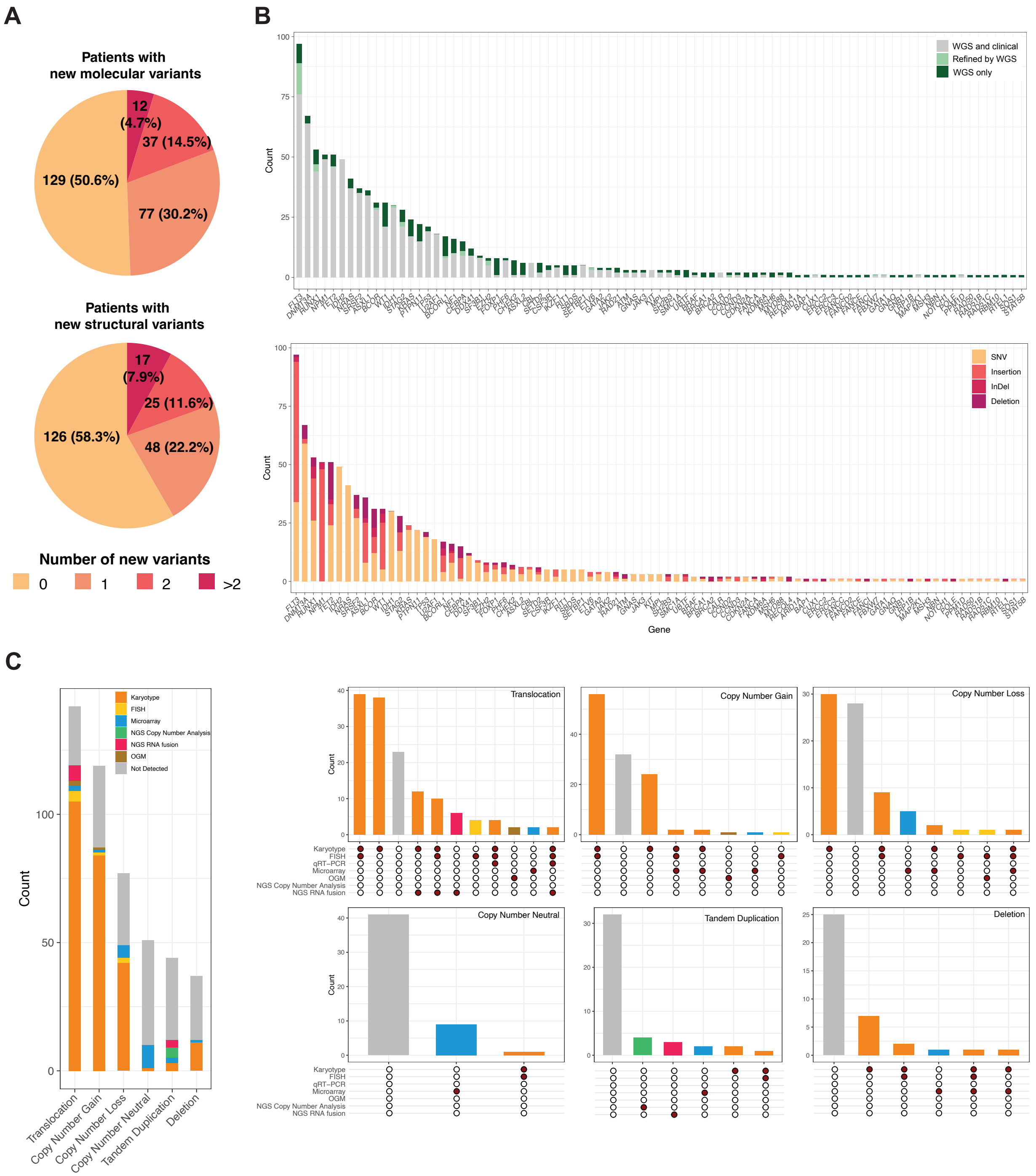


**Supplementary Fig. S6. Summary of variants detected by whole genome sequencing (WGS) and source of clinical tests for structural variants. (A)** The number and percentage of patients without or with novel genomic findings by WGS and not reported by diagnostic clinical testing (top: small variants; bottom: structural variants). The patients are also grouped by the number of new findings in each category. **(B)** Frequency of small variant calls by WGS compared with clinical testing (top) and variant types (bottom) on the gene level. **(C)** Bar plot of the number of structural variant calls from WGS by detailed variant type. Colors represent whether the variant was reported clinically, using the order from top to bottom in the legend. This means that if the variant was detected by multiple methods in clinical reporting, the color representing the first one from the list in the legend is used for that variant. Each sub-panel represents variants from one specific variant type and whether they were detected clinically. The dot grids show the combination of methods used clinically that detected the variants in the corresponding bar. See *Methods*, “Extraction of clinical testing results,” for details. Single nucleotide variant: SNV, Insertion Deletion: InDel, Fluorescence *In Situ* Hybridization (FISH), Next generation sequencing: NGS, Optical genome mapping: OGM, quantitative Real Time Polymerase Chain Reaction: qRT-PCR

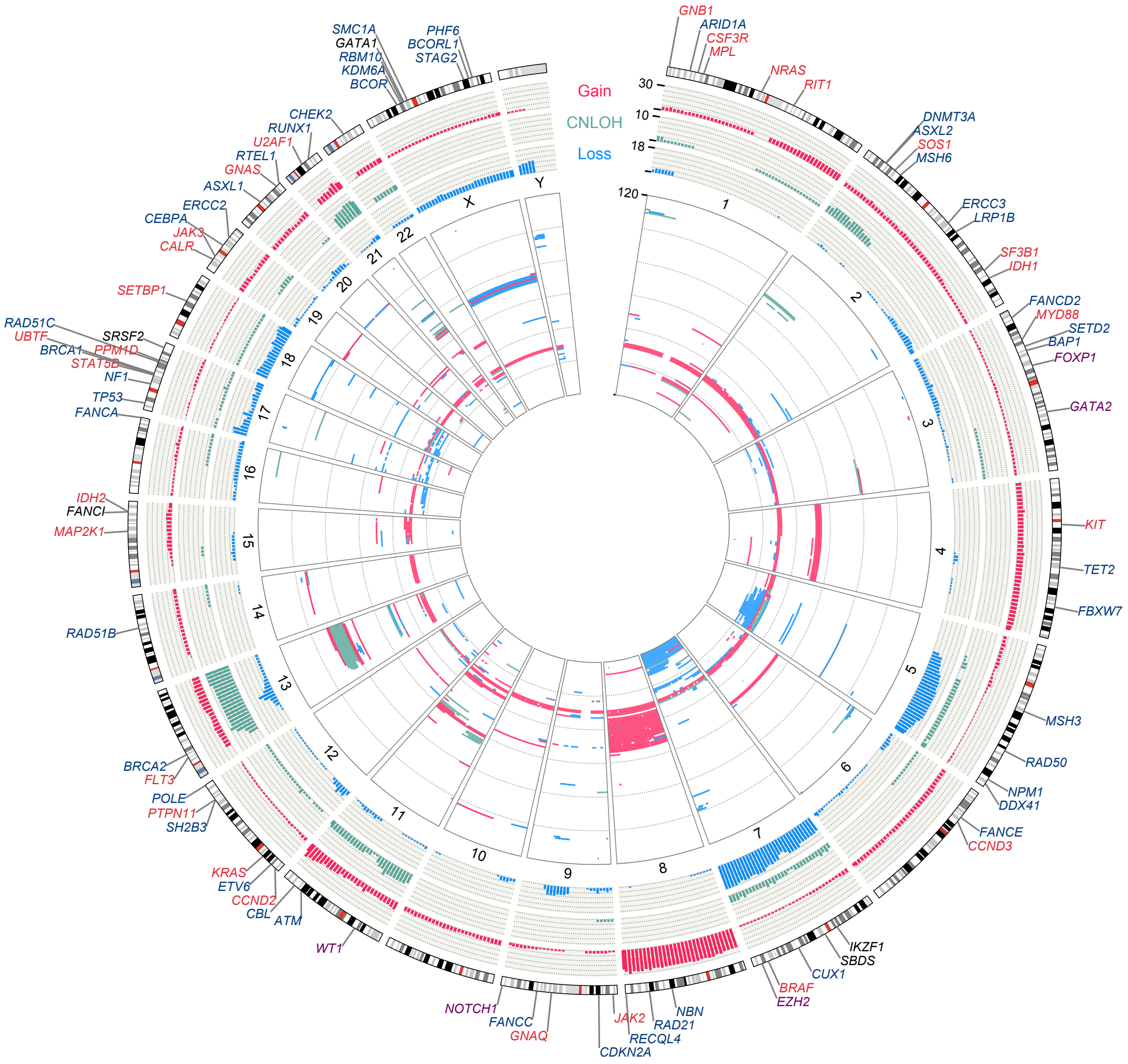


**Supplementary Fig. S7. Copy number variation (CNV) events in ATLAS cohort.** Rings outside the chromosome name are aggregated CNV events number in each of the gain, copy neutral loss of homozygosity (CNLOH), and Loss categories; and the cytoband plot of each chromosome. The heatmap ring inside the chromosome name is the patient level CNV events, color distinguished by the CNV categories (red: gain, loss: blue, CNLOH: green). Gene names annotated next to the cytobands are the location of AML and cancer-related genes. Red: oncogene; Dark Blue: tumor suppressor gene; Purple: oncogene and tumor suppressor gene; Grey: other.

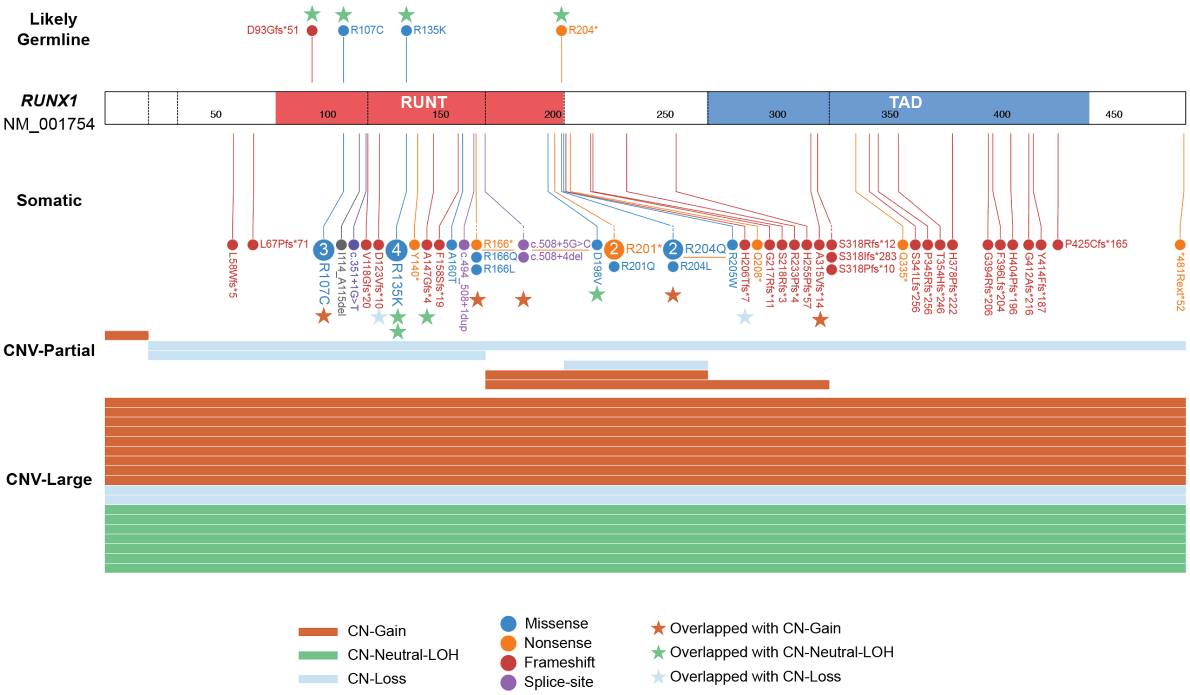


**Supplementary Fig. S8. *RUNX1* small variants and copy number variants identified in ATLAS cohort.**

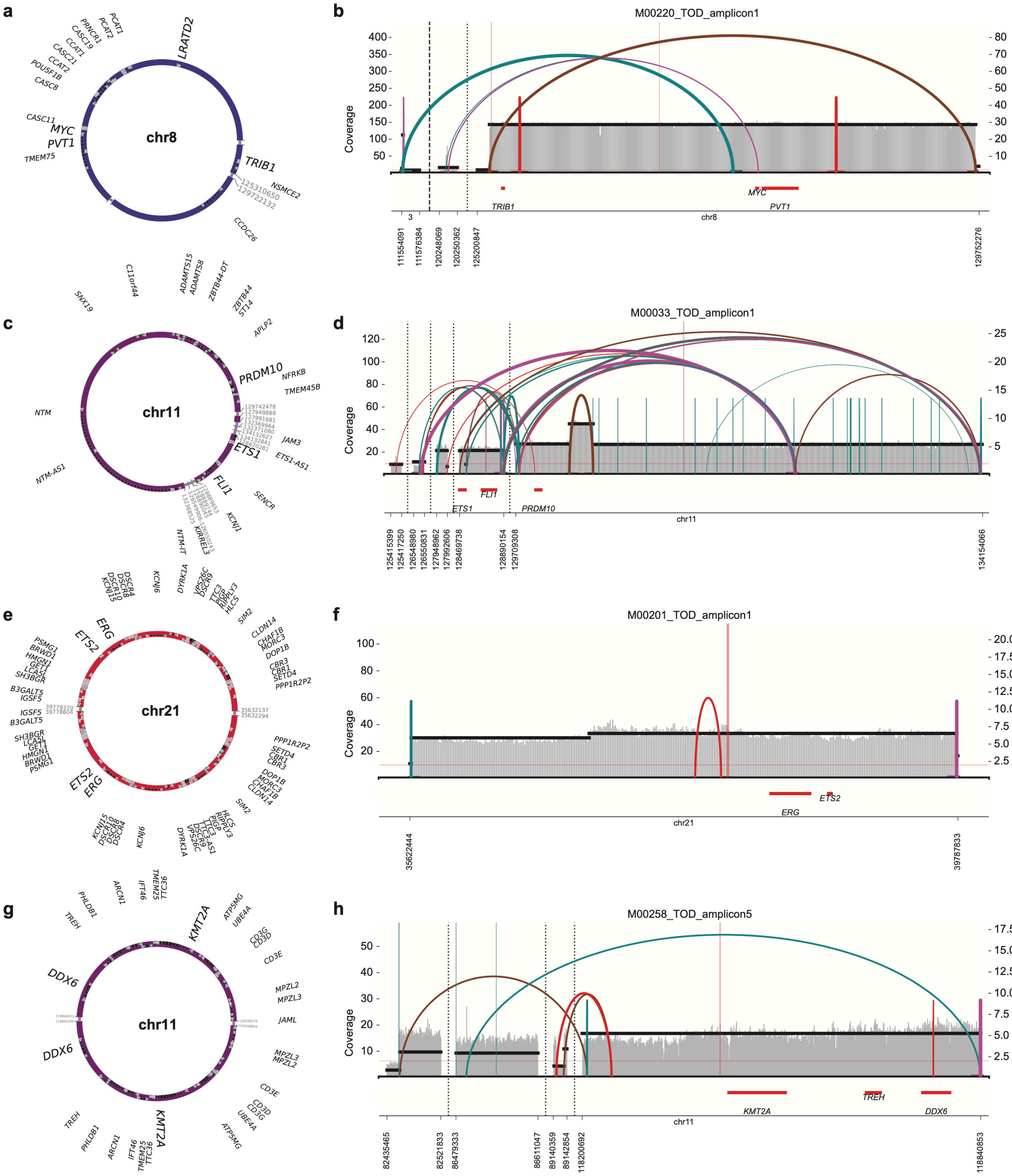


**Supplementary Fig. S9. Extrachromosomal DNA (ecDNA) in ATLAS cohort.**

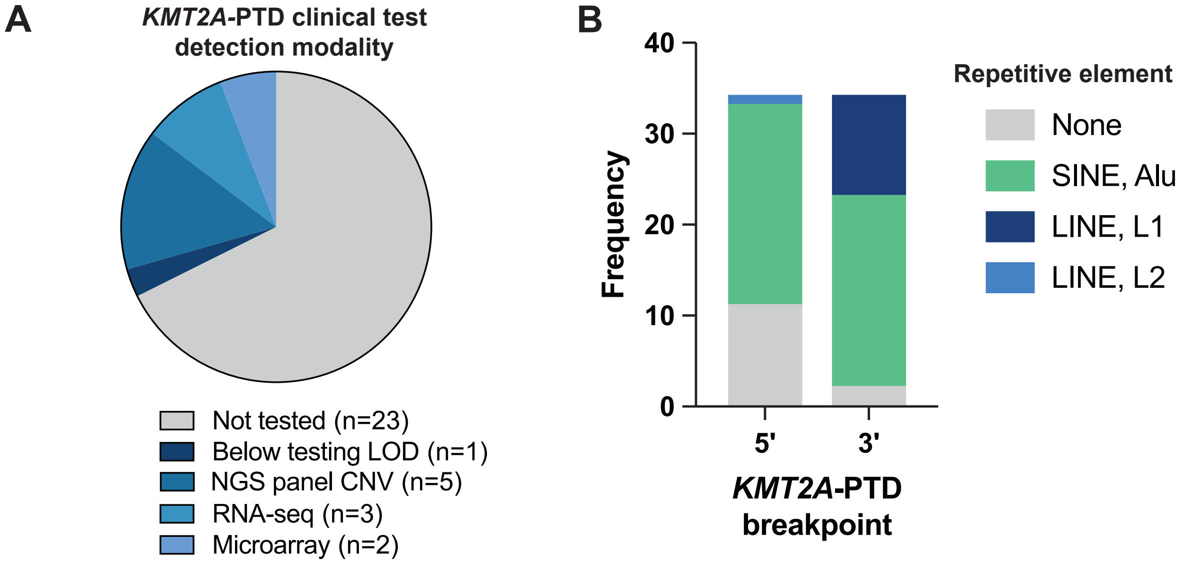


**C**

| Case ID | Gene | Variant type | Transcript | Exons | Introns | Note |
|---|---|---|---|---|---|---|
| MEA044 | *CBL* | MD | NM_005188.4 | 8 | 8 | *CBL* mutational hotspot: exon 8/9. Deletion of exon 8/9 reported in literature to result in loss of CBL activity in AML patients (PMID: 19276253). |
| MEA055 | *CBL* | MD | NM_005188.4 | 8-9 | 7-9 | |
| MEA119 | *ETV6* | MD | NM_001987.5 | 8 | 7 | Deletion includes exon 8 and 3'UTR of *ETV6*, minimally deleted region for *ETV6* includes 3'UTR region (PMID: 25213837, 9171997) |
| MEA132 | *ETV6* | MD | NM_001987.5 | 2-5 | 1-5 | Deletes HLH domain responsible for hetero- and homodimerization with other ETV6 proteins |
| MEA227 | *PHF6* | MD | NM_001015877.2 | 6-7 | 5-6 | Deletion of exons 5-6 destroy protein region known as necessary for tumor supressor function (PMID: 30917958) |
| MEA080 | *FOXP1* | MD | NM_001349338.3 | 6-11 | 5-11 | Deletion destroys dimerization domains of protein, results in early termination codon. |
| MEA194 | *CCDC26* | PTD | NR_130917.1 | 1 | 1 | Tandem duplications in *CCDC26* reported in literature to elevate MYC expression, duplicated region overlaps with common region reported (https://doi.org/10.1101/2021.03.14.435351). |
| MEA051 | *CCDC26* | PTD | NR_130917.1 | 1 | 1 | |
| MEA011 | *UBTF* | PTD | NM_014233.4 | 12-15 | 12-15 | Duplication begins in the middle of exon 12 through intron 15, including exon 13 critical region. One report of multi exon duplication with uncertain significance (PMID: 37085611) |
| MEA177 | *EZH2* | PTD | NM_004456.5 | 2-3 | 1-3 | Duplication impacts 5'UTR into start codon resulting in protein loss. |
| MEA246 | *UBTF* | ITD | NM_014233.4 | 13 | N/A | In-frame duplications occur within exon 13 of *UBTF*, consistent with literature (PMID: 37890156). |
| MEA045 | *UBTF* | ITD | NM_014233.4 | 13 | N/A | |
| MEA149 | *UBTF* | ITD | NM_014233.4 | 13 | N/A | |
| MEA148 | *IKZF1* | ITD | NM_006060.6 | 5 | N/A | Duplications in *IZKF1* have been reported in hematologic cancer (PMID: 37419244) |
| MEA150 | *IKZF1* | ITD | NM_006060.6 | N/A | 1 | |

**Supplementary Fig. S10. Additional details on other subgenic structural alterations detected by whole genome sequencing (WGS). (A)** Pie chart summarizing whether WGS-identified *KMT2A* partial tandem duplication (*KMT2A*-PTD) events were also detected by clinical genetic testing and the detection method used. **(B)** The location of *KMT2A*-PTD DNA breakpoints (5' or 3') within SINE/Alu and LINE repetitive elements was enriched, suggesting repetitive-sequence–mediated recombination. **(C)** Summary of other microdeletion (MD), PTD, and internal tandem duplication (ITD) observed by WGS. Next-generation sequencing: NGS, copy number variant: CNV.

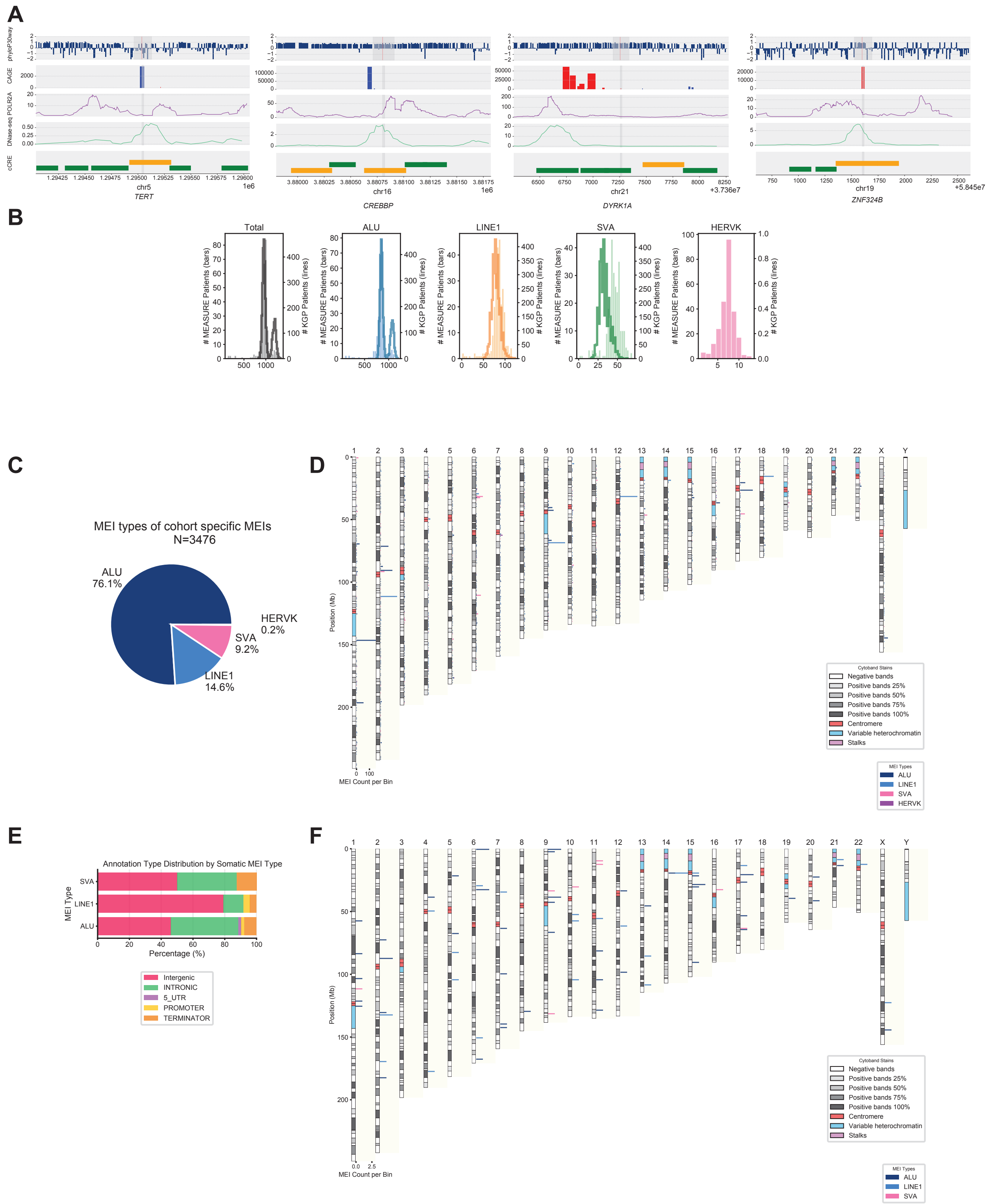


**Supplementary Fig. S11. Regulatory region variants and mobile element insertion (MEI). (A)** Tracks show multi-source genomic datasets around the variant locus (highlighted in gray). The phyloP30way conservation track indicates sequence conservation. The CAGE signal track marks transcription initiation activity around the variant. Red: positive strand signal; Blue: negative strand signal. POLR2A ChIP-seq track demonstrates active RNA polymerase II occupancy, and DNase-seq track shows open chromatin accessibility. The cCRE annotation identifies both PLS (orange) and pELS (green) elements in this region. **(B)** MEI event distribution of ATLAS cohort (bars) and 1000 Genome Project (1KGP) high-coverage WGS cohort (lines). ALU, LINE1, and SVA showed similar size profile; HERVK data were not available from 1KGP. **(C)** Cohort-specific MEI events dominated by ALU elements (76.1%), followed by LINE1 (14.6%), SVA (9.2%), and HERVK (0.2%). **(D)** Chromosomal distribution of MEI events detected in ATLAS cohort and not frequently seen in general population. The stacked histogram represents MEI density per 1-Mb bin, colored by MEI types. **(E)** Mobile element insertion location annotations, grouped by somatic MEI type. **(F)** Chromosomal distribution of somatic MEI events detected in ATLAS cohort. The stacked histogram represents MEI density per 1-Mb bin, colored by MEI types.

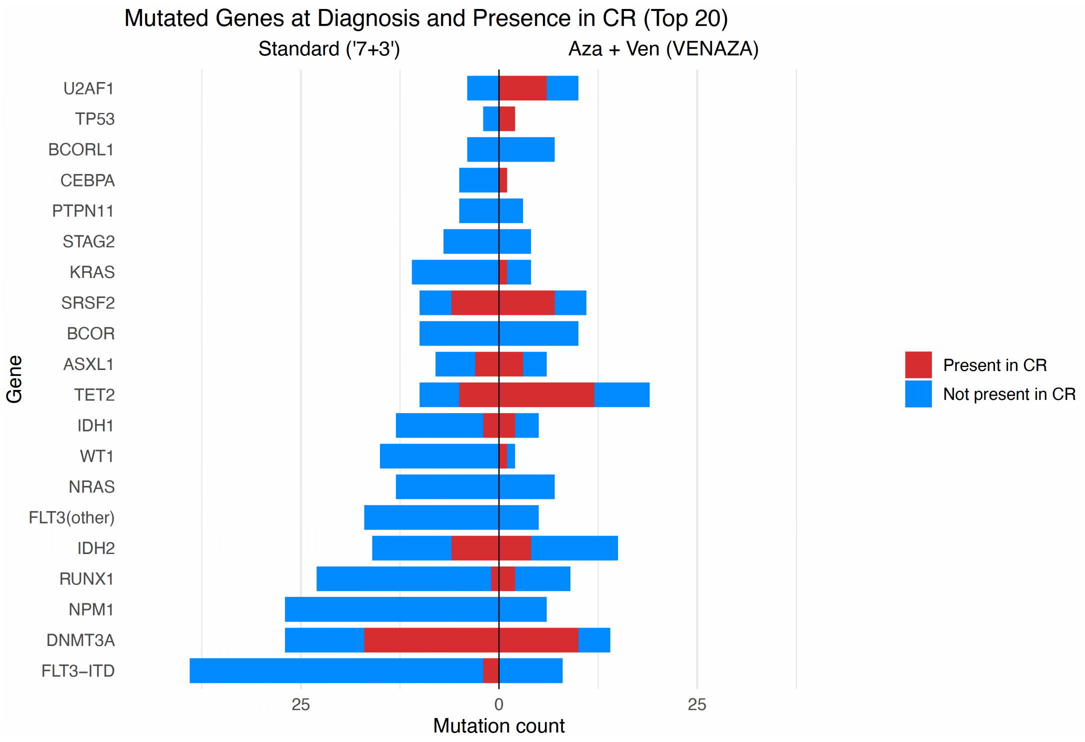


**Supplementary Fig. S12. Comparison of mutational persistence by induction regimens.** Bars on the left represent variants from patients with standard 7+3, and right from patients with azacitidine and venetoclax. Blue bars represent variants cleared at remission, and red bars indicate persistence.

**Supplementary Table S1. Patient level summary of whole-genome sequencing metrics for the ATLAS AML patient cohort.**

**Supplementary Table S2. Library level summary of whole-genome sequencing metrics for the ATLAS AML patient cohort.**

**Supplementary Table S3. Small variants detected by whole genome sequencing at the time of diagnosis.**

**Supplementary Table S4. Structural variants and copy number variants detected by whole genome sequencing at the time of diagnosis.**

**Supplementary Table S5. Summary of chromothripsis events detected by whole genome sequencing at the time of diagnosis.**

**Supplementary Table S6. Small variants detected by whole genome sequencing at the time of remission.**

**Supplementary Table S7. Structural variants and copy number variants detected by whole genome sequencing at the time of remission.**
